# Genome-wide insights into the shared genetic landscape between metabolic dysfunction-associated fatty liver disease and cardiovascular diseases

**DOI:** 10.1101/2024.11.10.24317047

**Authors:** Jun Qiao, Miaoran Chen, Minjing Chang, Wenjia Xie, Wenqi Ma, Tongtong Yang, Qianru Zhao, Kaixin Yao, Xichen Yang, Quan Yun, Jing Xiao, Xu He, Wen Su, Tao Xu, Yuliang Feng, Meixiao Zhan

## Abstract

**Background& Aims:** Multiple epidemiological studies have suggested an association between Metabolic dysfunction-associated fatty liver disease (MAFLD) and cardiovascular diseases (CVDs). However, the genetic components that are shared between the two remain unclear.

**Methods:** This genome-wide pleiotropic association study integrated comprehensive genome-wide association studies (GWAS) summary data from publicly available sources within European populations. It employed a range of genetic approaches to analyze the shared genetic architectures between MAFLD and six CVDs: atrial fibrillation (AF), coronary artery disease (CAD), venous thromboembolism (VTE), heart failure (HF), peripheral artery disease (PAD), and stroke. Initially, we examined the genetic correlation and overlap between these conditions. Subsequently, Mendelian Randomization (MR) analysis was conducted to investigate potential causal relationships. Finally, we explored horizontal pleiotropy at the levels of single nucleotide polymorphisms (SNPs), genes, and biological pathways to further elucidate the shared genetic mechanisms underlying.

**Results:** We observed significant genetic associations between MAFLD and four CVDs, including CAD, HF, PAD, and VTE. However, we noted extensive genetic overlap in all but MAFLD-AF. MR analysis established causal relationships from MAFLD to both AF and PAD. Regarding horizontal pleiotropy, 49 pleiotropic loci were identified at the SNP level with functional annotations, 13 demonstrating strong evidence of colocalization. At the gene level, 14 unique pleiotropic genes were found, with SAMM50 (located at 22q13.31) being particularly notable. Further pathway enrichment analysis indicated that these genes significantly contribute to the pathway of establishment of protein localization to membrane, highlighting their pivotal role in the pathophysiology of both MAFLD and CVD.

**Conclusions:** In all, our research proved the shared genetic architectures and mechanisms between MAFLD and CVD and elucidated their shared genetic etiology and biological mechanisms.

**Impact and implications:** Metabolic dysfunction-associated fatty liver disease (MAFLD) has reached a prevalence of 25-30% worldwide and has emerged as a global leading cause of liver-related morbidity and mortality. Studies have shown that people with MAFLD have a higher risk of cardiovascular disease (CVD) than the general population and there is currently no effective drug to treat the comorbidity of the two, which imposes a burden on the socioeconomic situation and the adverse effects are still rising. Therefore, it is critical to understand how MAFLD affects CVD. Our study provides unique insights into the mechanisms of comorbidity between MAFLD and CVD. The increasing number of complications has prompted us to explore new treatment options, so our study has important clinical significance.

**Graphical abstract:** 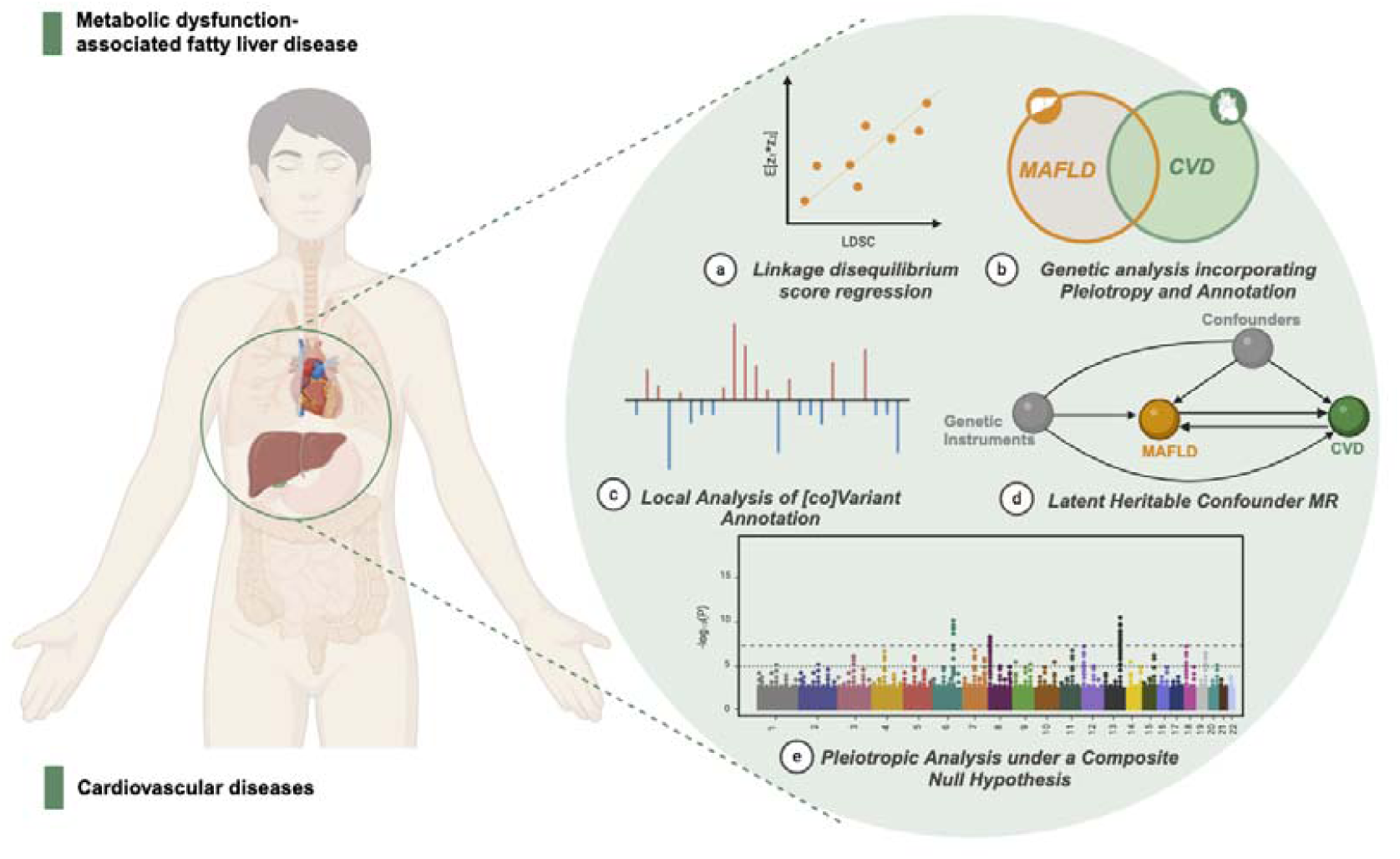

**Highlights:**

- The first comprehensive and systematic study to explore the common genetic components between MAFLD and CVD.
- MAFLD and CVDs share genetic architectures and mechanisms.
- Genetically predicted MAFLD increases the risk of AF and PAD.
- The effects of SAMM50 (located at 22q13.31) on lipid metabolism support the comorbidity of MAFLD and CVDs.
- The localization of lipid droplet related contact site proteins to the membrane plays a key role in the comorbidity of MAFLD and CVD.

## Introduction

Non-alcoholic fatty liver disease (NAFLD) encompasses a range of liver disorders, from simple steatosis to non-alcoholic steatohepatitis, fibrosis, and cirrhosis, typically arising without significant alcohol consumption. According to a recent consensus statement by an international panel of experts, Metabolic dysfunction-Associated fatty liver disease (MAFLD) has been proposed as a more appropriate and comprehensive term to replace NAFLD, better reflecting its association with known metabolic dysfunctions^1,2^. MAFLD has emerged as a global leading cause of liver-related morbidity and mortality^3^. While liver-related complications significantly elevate mortality rates, cardiovascular diseases (CVDs) remain the primary cause of death in patients with MAFLD^4^. Clinically, individuals with MAFLD often exhibit elevated triglyceride levels and an increased concentration of residual lipoprotein particles, thereby elevating their risk for CVDs^5^. Lipoproteins containing apolipoprotein C3 (ApoC3) have been shown to activate the IL(interleukin)-1 to IL-6 to CRP (C-Reactive Protein) inflammatory pathway, which is implicated in the development of coronary artery disease (CAD), venous thromboembolism (VTE), Stroke, and vascular inflammation^6^. A meta-analysis of 16 observational studies, encompassing 34,043 patients over a median follow-up of 6.9 years, reported a 64% increased risk of fatal and/or non-fatal CVD events in patients with MAFLD compared to those without^7^. The strong epidemiological evidence underscores the association between MAFLD and an escalated risk of CVDs, which might suggest increased genetic liability to CVD in subgroups of patients with MAFLD.

Enhanced comprehension of the shared genetic foundations between MAFLD and various CVDs could offer valuable insights into these conditions. Recent large-scale genome-wide association studies (GWAS) have uncovered numerous risk loci linked to both MAFLD and CVDs, identifying several shared risk loci, notably at 19p13.11 (*TM6SF2*)^8^ and 22q13.31 (*PNPLA3*)^9^. Increasing evidence suggests genetic overlap between MAFLD and CVDs. For instance, recent data suggested weak to moderate positive genetic correlations between MAFLD and CAD, heart failure (HF), and Hypertension, a major risk factor for CVDs^10^. Genetic correlation provides a summary measure of the correlation across all single-nucleotide polymorphism (SNP) effect sizes. However, it cannot differentiate between genetic overlap with a combination of concordant and discordant effects and a complete absence of genetic overlap, frequently resulting in an estimate close to 0 in both scenarios^11^. While genetic correlation can provide evidence for overall genomic similarity between traits, it does not provide information for considering biological plausibility or inferring potential causal relationships^12^. Previous Mendelian randomization(MR) analyses investigating the link between MAFLD and CVDs have shown that the causal relationship was not consistently robust due to possible bias by the presence of horizontal pleiotropy or sample overlap^13–15^. Despite these insights, a significant portion of the genetic underpinnings of MAFLD and CVDs remain elusive. Moreover, individual loci associated with both conditions have not been systematically analyzed, an examination of which could clarify the impact of different risk loci on the comorbidities of MAFLD and CVDs, as well as identify biological pathways offering therapeutic promise.

This study aims to uncover their genetic overlap beyond the genetic correlation between MAFLD and six major CVDs—atrial fibrillation (AF), CAD, VTE, HF, peripheral artery disease (PAD), and Stroke—by leveraging unprecedentedly large GWAS summary data from European ancestry. We applied the genetic analysis incorporating pleiotropy and annotation (GPA), which estimates the total number of unique and shared genetic variants between pairs of traits. The relevance of mixed effects has been further emphasized by Local Analysis of [co]Variant Annotation (LAVA), which calculates local genetic correlations across the genome, even in the presence of minimal *r_g_*. Given that GPA and LAVA quantify total genetic overlap but cannot pinpoint shared genomic loci, we subsequently utilized the Pleiotropic Analysis under a Composite Null Hypothesis (PLACO) method to discover loci jointly associated with MAFLD and each CVD beyond genome-wide significance, a critical step for gaining biological insights. Additionally, we also utilize Latent Heritable Confounder Mendelian randomization (LHC-MR) analyses to explore potential causal relationships between MAFLD and CVDs, considering sample overlap, bidirectional causal associations, and unobserved heritable confounders. Together, these methods augment the insights gained from genetic correlation analyses, shedding light on the distinct and common genetic frameworks underpinning MAFLD and CVDs, with implications for how we conceptualize genetic risk for the comorbidity between MAFLD and CVDs.

## Materials and methods

### Data sources and quality control

Genetic associations with MAFLD were derived from the largest genome-wide meta-analysis to date, involving 8,434 cases and 770,180 controls from four European cohorts: the Electronic Medical Records and Genomics (eMERGE) Network, the UK Biobank (UKB), the Estonian Biobank (EstBB), and the FinnGen Consortium, of which the diagnosis was determined based on the electronic health records of all participants^16^. To ensure that the sample size is large enough to produce reliable results, we use GWAS summary data from studies with sample sizes greater than 50,000. Accordingly, our study encompassed six major CVDs, including AF, CAD, VTE, HF, PAD, and Stroke. Specifically, GWAS summary statistics for AF were retrieved from a recent meta-analysis of six cohorts, including the Health Study Nord-Trøndelag (HUNT), deCODE, Michigan Genome Initiative (MGI), DiscovEHR, UKB, and AFGen consortium, comprising 60,620 cases and 970,216 controls of European ancestry^17^. GWAS summary statistics for CAD were extracted from a genome-wide meta-analysis by the CARDIoGRAMplusC4D and UKB consortium, including 181,522 cases and 984,168 controls of European ancestry^18^. GWAS summary statistics for VTE were collected from the largest meta-analysis of seven cohorts to date, including 81,190 cases and 1,419,671 controls of European ancestry^19^. GWAS summary data for HF were derived from the Heart Failure Molecular Epidemiology for Therapeutic Targets (HERMES) consortium, entailing 47,309 cases and 930,014 controls^20^. GWAS summary data for PAD came from the largest meta-analysis conducted on data from 11 independent cohorts, totaling 12,086 cases and 499,548 controls^21^. GWAS summary data for Stroke were obtained from the GIGASTROKE consortium, which included 73,652 cases and 1,234,808 controls^22^. Detailed information about these GWAS summary data and their original publication sources is available in Supplementary Table 1.

Quality control was carried out strictly for GWAS summary data per the following steps before further analysis:(i) alignment with the hg19 genome assembly, and the 1000 Genomes Project v3 Europeans was used as the reference^23^; (ii) only including autosomal SNPs; (iii) SNPs with duplicate entries or missing rsIDs were eliminated; (iv) SNPs with a minor allele frequency (MAF) value of 0.01 or less were excluded.

To ensure robust results, we normalized the GWAS summary data for all phenotypic SNPs, yielding 6,479,654 common SNPs across all diseases.

### Assessing the genetic correlates of MAFLD and CVDs

We employed cross-trait linkage disequilibrium score regression (LDSC) analyses to assess the genetic correlation (*r_g_*) between MAFLD and six major CVDs^24^. LDSC, an effective method for evaluating genetic correlations across the genome, utilizes the linkage disequilibrium (LD) structure to estimate individual SNP effect sizes from GWAS summary statistics^25^. The method involves regression of these GWAS summary statistics against LD scores derived from the European 1000 Genomes Project, minimizing biases related to polygenicity, sample overlap, and population stratification^24^. Due to its complexity, the major histocompatibility complex (MHC) region was excluded from our analysis. First, univariate LDSC was conducted to determine the SNP-based heritability (*h^2^_SNP_*) for MAFLD and each CVD. Subsequently, bivariate LDSC estimated the *r_g_*between MAFLD and CVDs by performing a weighted linear regression on the product of Z-scores for MAFLD and CVDs against the LD scores for all available genetic variants. This approach provides an unbiased estimate of *r_g_*and reliably measures genetic correlations even in overlapping samples between GWAS datasets. Genetic correlations rang from -1 to +1, with values closer to the extremes indicating stronger influences, and negative values suggest opposite-direction effects, while positive values indicate the same direction. *P*-values below a Bonferroni-corrected significance threshold (*P* = 0.05 / 6 = 8.3×10^-3^) were considered statistically significant.

To identify tissues closely associated with genetic susceptibility SNPs for MAFLD and CVDs, we used LDSC applied to specifically expressed genes (LDSC-SEG)^26^. This method combines aggregate GWAS summary statistics with tissue-specific gene expression data for targeted tissue enrichment analysis^26^. Our analysis incorporated various gene sets, including multi-tissue gene expression data from the GTEx project (v8) and chromatin information from the Roadmap Epigenomics and ENCODE initiatives^26,27^. Using the baseline model and comprehensive gene sets, we prioritized genes from GTEx based on computed t-statistics, focusing on the top 10% of specifically expressed candidate genes that exhibited the highest t-statistics. The P-value from the regression coefficient’s z-score was used to determine the significance coefficient, and a false discovery rate (FDR) of less than 0.05 was applied to assess the significance of enriched SNPs in specific tissues.

### Estimating genetic overlap between MAFLD and CVDs

To address the “missing heritability” issue in complex traits, the Genetic Analysis incorporating Pleiotropy and Annotation (GPA) was specifically developed. This tool was specifically designed to overcome the limitations posed by the small proportion of genetic variants identified through standard approaches that account for only a minimal part of the expected genetic contribution^28^. GPA achieves this by integrating diverse genomic data and annotation information, thereby enhancing the prioritization of GWAS results and providing a more comprehensive estimation of genetic overlap. Notably, in the absence of annotation data, GPA can infer genetic overlap using p-value interactions of the corresponding phenotypes^28^. GPA is based on key assumptions regarding p-value distributions: p-values from null SNPs are expected to adhere to a uniform distribution, reflecting no association, while p-values from non-null SNPs are presumed to follow a beta distribution, indicating potential associations. Within this framework, GPA categorizes SNPs into four groups based on their p-value characteristics: π00 suggests no association with either trait; π01 denotes an association exclusively with the first trait; π10 signifies an association solely with the second trait; and π11 represents an association with both traits. This classification can estimate π11 / (π10 +π11 +π11), an important ratio indicator that represents the proportion of SNPS shared by two traits, suggesting the extent to which a common biological pathway may be shared between the two traits^28^. Subsequently, the likelihood ratio test (LRT) was used to assess the statistical significance of the observed genetic overlap^28^. Given the complex interrelations among SNPs, GPA estimates are susceptible to biases from LD. We used PLINK1.9 for LD pruning to address this, drawing on data from the 1000 Genomes Project Phase 3^23^ for European ancestry. We also applied a Bonferroni-corrected significance threshold for multiple comparisons, set at *P* = 0.05 / number of trait pairs = 0.05 / 6 = 8.3×10^-3^.

### Calculating local genetic correlations between MAFLD and CVDs

LAVA facilitates the assessment of local genetic correlations by partitioning the genome into smaller regions, which enables detailed insights into the influence of genetic variants on various traits at a regional level, rather than across the entire genome^29^. LAVA analyses genetic correlations within different genomic segments, using the regions delineated by Werme et al. as autosomal LD blocks^29^, characterized by minimal connectivity between blocks. The genome was partitioned into 2,495 semi-independent regions, each approximately 1 Mb, using genotype data from the 1000 Genomes Project Phase 3 European ancestry as the LD reference panel and MHC region (chr 6: 25-35 Mb) was excluded. In conducting the genetic association study, we first performed a preliminary univariate analysis using LAVA to estimate the local heritability of MAFLD and CVDs. With this step, we could identify regions that showed significant signals in the local genetic analysis. We then performed bivariate LAVA analyses of these regions to explore regional genetic associations between MAFLD and CVDs. To ensure the statistical significance of the results, the FDR method was employed to calculate the adjusted p-value. FDR less than 0.05 was considered the genetic correlation of these regions to be statistically significant.

### Evaluate the causal relationship between MAFLD and CVDs

The genome-wide and local genetic correlations provide insights into the shared genetic basis between trait pairs, but these correlations do not imply causality in either direction. To overcome these limitations, linkage disequilibrium heterogeneity causal Mendelian randomization (LHC-MR) provides a more refined approach to assess bidirectional causality while minimizing confounding effects^30^. This method extends traditional MR by addressing limitations of the standard two-sample MR approach, such as potential sample overlap, under-utilization of genome-wide markers, and the need to understand exposure-outcome relationships. LHC-MR assesses bidirectional causality by categorizing associations between exposure and outcome into four categories: (i) causal effect of exposure on outcome, (ii) causal effect of outcome on exposure, (iii) effect of confounders whose exposure affects the outcome process (vertical pleiotropy), and (iv) effect of confounders whose exposure affects the outcome independently (horizontal pleiotropy^30^). Unlike traditional MR, LHC-MR utilizes all genome-wide genetic variation rather than only genome-wide significant variation^30^. To ensure the stability of the results, the Bonferroni correction significance threshold was set at 4.17 × 10^-3^ (*P* = 0.05/number of trait pairs/number of tests). When *P_axy_* < 4.17×10^-3^ and *P_ayx_* > 0.05, it suggests a one-way causal relationship from MAFLD to CVD; conversely, when *P_ayx_* < 4.17×10^-3^ and *P_axy_* > 0.05, it implies a one-way causal relationship from CVD to MAFLD. However, if both *P_axy_*and *P_ayx_*, exceeding the significance threshold, it indicates bidirectional causality between MAFLD and CVD. We used traditional two-sample MR techniques for sensitivity analysis, including Inverse Variance Weighting (IVW), MR Egger, Weighted Median, Simple Mode, and Weighted Mode. *P*-values below the 0.05 threshold were considered statistically significant.

### Identification of pleiotropic loci between MAFLD and CVDs

To unravel the shared genetic mechanisms between MAFLD and six major CVDs, our study employs the PLACO, extending pleiotropic analysis to the SNP level^31^. PLACO utilizes the summary statistic from GWAS to calculate the test statistic for the product of the Z-statistics for each SNP in the two trait analyses. This calculation is based on the assumption of a mixed distribution, which allows for the assessment of SNP associations across traits. Using this assumption, PLACO can rigorously analyze individual SNPs by pairwise Z-statistics, categorizing the null hypothesis of pleiotropy into three distinct and mutually exclusive scenarios: (i) H00, indicating no association of the SNP with either trait; (ii) H10, signifying an association with the first trait but not the second; and (iii) H01, representing an association with the second trait but not the first. The rejection of the composite null hypothesis, which encompasses all three scenarios, indicates horizontal pleiotropy, suggesting that the SNP influences both traits through identical mechanisms. We applied a stringent significance threshold of 5×10^-8^ to identify SNPs exhibiting statistically significant pleiotropy.

### Functional annotation based on pleiotropic loci

To identify and conduct independent genomic loci between MAFLD and six major CVDs, we utilized the Functional Mapping and Annotation (FUMA) platform^32^. FUMA, an online tool designed to enhance the interpretability of GWAS findings, performs functional annotation of pleiotropic SNPs revealed by PLACO analysis. Lead SNPs are merged to delineate distinct genomic loci in LD blocks within 500 kb of lead SNPs by FUMA, utilizing default parameters and LD data from the 1000 Genomes Project Phase 3 European cohort. Lead SNP was further selected by r^2^ < 0.1 for minimal LD in independent significant SNPs (genome-wide significance *P* < 5×10^-8^ and r^2^ < 0.6), indicating higher independence from adjacent genetic variations. Despite these regions potentially containing several lead SNPs, the Top Lead SNP is identified with the lowest P-value within a given region. Locus was deemed novel to MAFLD and CVD if they did not coincide with the loci previously reported in the original GWAS.

Functional annotations for Top Lead SNPs, provided by FUMA, include Combined Annotation-Dependent Depletion (CADD^33^) scores, regulatory function probability scores via RegulomeDB (RDB^34^), and potential effects on gene function as annotated by ANNOVAR^35^. CADD scores, which assess the harmful impact of SNPs, are utilized with a threshold: a score greater than 12.37 is considered likely to be harmful, reflecting the SNP’s potential to affect biological functions adversely. RDB scores, ranging from 1 to 7, indicate the likelihood of an SNP being located in a regulatory function area, with lower scores denoting higher functional significance. Lead SNPs were annotated using ANNOVAR to evaluate their proximity to genes and potential impacts on gene functions, thus providing insights into how these variants might influence genetic pathways or disease mechanisms. For the identification of putative causal genes, SNPs were mapped using two approaches: positional mapping within a 10-kb window around the SNP and eQTL mapping.

### Colocalization analysis

After annotating pleiotropic loci using FUMA, we analyzed colocalization with the “COLOC” R package to identify potential shared causal variants across trait pairs within each locus. COLOC analysis employs Bayes factors to evaluate the likelihood of shared causal variation between two sets of trait-associated loci, calculating posterior probabilities (PP) for five mutually exclusive hypotheses at each locus: (i) no SNP is associated with either trait (H0); (ii) a causal SNP is associated only with the first trait (H1); (iii) a causal SNP is associated only with the second trait (H2); (iv) each trait is influenced by independent, distinct causal SNPs (H3); and (v) a single SNP acts as a causal variant for both traits (H4). Our analysis primarily focuses on hypothesis H4, which posits a shared causal variant for both traits. Strong evidence of colocalization at a genomic locus is indicated if the posterior probability for shared causal variants (PPH4) exceeds 0.70. The SNP with the highest PPH4 within these loci is identified as the candidate causal variant.

### Gene-level annotation analysis

To pinpoint genes that exhibit pleiotropic effects within genomic loci, a Multi-marker Analysis of GenoMic Annotation (MAGMA) was performed based on the results of PLACO and individual GWAS^36^. MAGMA, a tool rooted in multivariable regression models, computes the significance of gene-trait associations by incorporating principal component analysis. It then employs the F-test to ascertain p-values for each gene, considering factors such as gene size, SNP count per gene, and LD among the markers. Consistency in gene-based testing was ensured by defining gene boundaries, which encompassed regions extending ±10 kb from the termini of each gene. We obtained MAGMA gene IDs and location data for 17,636 protein-coding genes, aligning SNP locations with Human Gene Build 37 (GRCh37/hg19). The MHC region (chr6: 25-35 Mb) was excluded from MAGMA’s gene-based analysis. Significant gene associations were identified using a stringent Bonferroni-corrected threshold set at *P* = 4.73×10^-7^ (0.05 / 17,636 / 6), accounting for the number of protein-coding genes and trait pairs analyzed.

Recognizing that MAGMA links SNPs to genes based primarily on physical proximity, we extended our analysis to include the eQTL-informed MAGMA (e-MAGMA)^37^. The e-MAGMA utilizes tissue-specific eQTL data to assign risk variants to presumptive genes, thereby converting genome-wide association summary statistics into gene-specific metrics. This method leverages tissue-specific eQTL data from the GTEx project to account for long-range regulatory effects, enhancing the interpretation of GWAS association signals. MHC region (chr6: 25-35 Mb) was excluded in e-MAGMA analysis results as well as MAGMA to avoid confounding due to complex LD patterns. Our study utilizes the GTEx v8 comprehensive database containing eQTL data from 47 different tissues, based on PLACO results, to delve into gene-disease associations at a fine-grained tissue level, and to gain a more detailed understanding of the tissue-specific genetic architecture behind the relevant diseases. To minimize confounding factors associated with broad tissue analyses, we refined our approach by selecting a subset of tissues based on insights from LDSC-SEG results and previous research. Specifically, our focused analysis included ten tissues: two types of adipose tissue (subcutaneous and visceral omental fat), three arterial tissues (aorta, coronary, and tibial arteries), two cardiac tissues (left ventricle and atrial appendage), as well as liver, EBV-transformed lymphocytes, and whole blood. Using the 1000 Genomes Project v3 European samples as the LD reference, we calculated tissue-specific p-values for each gene across the selected tissues. We applied a Bonferroni correction to determine the significance of these p-values, accounting for the number of tissue-specific protein-coding genes and trait pairs examined. For instance, the significance threshold for adipose subcutaneous tissue was set at *P* = 0.05 / 7,560 / 6 = 1.10×10^-6^.

To further explore tissue-specific gene-trait associations for MAFLD and CVDs, we conducted a transcriptome-wide association study (TWAS) using single-trait GWAS results^38^. Employing the FUSION framework, TWAS integrates GWAS summary statistics with eQTL data from the GTEx v8 dataset to assess how variations in gene expression across specific tissues influence phenotypes. We used pre-computed gene expression weight references from the tissues analyzed in our e-MAGMA study. LD pruning was performed during the third phase of the 1000 Genomes Project using the LD structures of the European subpopulation. Significance thresholds were adjusted using tissue-specific Bonferroni corrections to control for multiple tests.

### Pathway level analyses

We performed functional enrichment analysis on the overlapped gene lists significantly identified by MAGMA and e-MAGMA analyses using the ToppGene functional annotation tool (ToppFun)^39^. ToppGene Suite is a comprehensive portal for gene list enrichment analysis and candidate gene prioritization. The ToppFun application provides a variety of annotation data types, including transcriptome, proteome, regulatory elements (such as transcription factor binding sites (TFBS) and microRNA), ontologies (such as GO and Pathway), phenotypes (including human diseases and mouse phenotypes), pharmacometrics (drug-gene associations), and literature co-citations. Additionally, ToppFun prioritizes candidate genes based on their functional similarity to ensure rigorous and thorough analysis. Significant GO terms were identified with an FDR threshold of 0.05.

## Results

### SNP-based heritability and genetic correlations between MAFLD and CVDs

After harmonizing and filtering SNPs common across GWAS summary statistics, LDSC was utilized to estimate the *h^2^_SNP_* and the genome-wide genetic correlations (*r_g_*) between MAFLD and CVDs. Univariate LDSC analysis indicated that the *h*^2^ of MAFLD was 0.002 (SE = 0.001). In comparison, the *h^2^_SNP_* of CVDs ranged from 0.008 to 0.032, with CAD exhibiting the highest heritability (*h*^2^ = 0.032, SE = 1.90×10^-3^) and HF the lowest (*h^2^_SNP_* = 0.008, SE = 3.01×10^-3^). Notably, the heritability of MAFLD was lower than that of any individual CVD. Bivariate LDSC revealed that all trait pairs exhibited positive genetic correlations under a relaxed significance threshold (*P* < 0.05). However, four trait pairs maintained statistical significance after applying a Bonferroni correction for multiple comparisons (*P* < 0.05 / 6 = 8.33×10^-3^). These findings highlight the subtle genetic interactions between MAFLD and CVD, which underscores the potential for shared genetic pathways to influence these complex diseases.

### Genetic overlap analysis

The *r_g_* is a vital metric that reflects the genetic relationship between trait pairs, providing insights into their shared genetic architecture. Importantly, a *r_g_* value close to zero does not necessarily signify the absence of a genetic association between traits. This apparent discrepancy can arise from various factors, such as the mixed effects of homozygosity and reverse effects, or a lack of genetic overlap, which may mask the true genetic relationships and lead to misleadingly low *r_g_* values. Recognizing this limitation highlights a “missing dimension” in our understanding of genetic connections between phenotypes. To enhance our understanding of genetic interactions and address these challenges, advanced statistical methods, including GPA and LAVA, have been employed by us.

The intricacies of the genetic relationships observed between MAFLD and CVDs, particularly the surprising contrast between significant genetic overlap and weak correlation in some cases, underscore the complexity of genetic interactions and the importance of considering additional analytical dimensions. In our analysis of six trait pairs, the MAFLD-AF pair was the only one that failed to meet the Bonferroni correction threshold, indicating a lack of significant genetic overlap. Conversely, the MAFLD-CAD pair demonstrated the most significant genetic overlap, evidenced by a P-value of 7.12×10^-31^, aligning with LDSC results, which revealed the highest genetic correlation (*r_g_* = 0.627, *P* = 5.18×10^-6^). Although a significant genetic overlap was identified between MAFLD and Stroke (*P* = 6.96×10^-6^), the minimal PAR value (PAR = 7.00×10^-4^) highlights a small proportion of pleiotropic SNPs. Furthermore, the weak genetic correlation between MAFLD and Stroke (*r_g_* = 0.627, *P* = 3.98×10^-2^) suggests that the presence of multiple closely linked SNPs with independent or heterogeneous impacts may obscure consistent effects, resulting in a correlation that falls short of higher statistical confidence thresholds. This phenomenon, where broad genetic overlap coexists with weak genetic correlation, implies that mixed effects may mask the true genome-wide genetic correlation.

### Regional genetic correlation analysis

LAVA evaluates genetic correlations within specific genomic regions and identifies subtle genetic effects that might elude detection by LDSC. This capability is vital, as genetic influences often vary across the genome, and the correlations between traits can differ markedly by region. First, a univariate LAVA analysis was conducted by us with a strict threshold of *P* < 1×10^−4^ to identify regions with strong genetic signals, resulting in the identification of 2,336 significant regions. These regions were then formed into 77 bivariate test sets for more detailed analysis. In the subsequent bivariate LAVA analysis, we applied a looser significance threshold (*P* < 0.05), identifying approximately 22 genomic regions showing genetic signals associated with all trait pairs. Notably, our LAVA analysis revealed five key genetic regions associated with MAFLD and stroke, including three negatively and two positively correlated regions. This discovery of substantial genetic overlap starkly contrasts the lack of significant genome-wide associations found using LDSC. This discrepancy between the LAVA and LDSC results underscores the complex genetic architecture, suggesting that regions with both positive and negative correlations may lead to a heterogeneous distribution of genetic effects. Such complexity might mask genetic signals when assessed on a genome-wide scale using LDSC. We then applied a stricter threshold of *P* < 6.49×10−4 to account for multiple comparisons. This led to the identification of a significant region on chromosome 8, spanning from 125,453,321 to 126,766,827, showing a strong correlation between MAFLD and CAD (*r_g_s*=0.509, *P* =3.05×10^-6^), aligns with the positive genetic correlation observed across the genome by LDSC.

### Causal relationship linking MAFLD and CVDs

The observed genetic overlap between MAFLD and CVDs extends beyond mere correlation, suggesting the presence of pleiotropy. However, determining whether this pleiotropy is horizontal or vertical remains unresolved. To clarify this, we utilized the LHC-MR approach, which sheds light on the bidirectional causal relationships between MAFLD and CVD through vertical pleiotropy. Our findings revealed a significant positive causal effect of MAFLD on both AF and PAD, with odds ratios (OR) of 1.11 (*P* = 2.36×10^-3^) and 1.375 (*P* = 1.39×10^-4^), respectively. These results underscore MAFLD as a critical risk factor in the onset and development of both AF and PAD, aligning with previous research that highlights MAFLD’s role in increasing the prevalence of these disorders. Notably, we found no evidence to suggest reverse causality in these relationships.

### Shared variants and functional annotation of MAFLDs and CVDs

To further dissect the complex genetic mechanisms between MAFLD and CVDs, we utilized PLACO to identify significant pleiotropic variants indicative of horizontal pleiotropy. This inquiry, through its meticulous analysis, unveiled a trove of 3,438 SNPs exhibiting significant pleiotropic influences. Subsequently, these SNPs underwent a process of annotation, facilitated by the FUMA tool, which meticulously delineated 49 distinct loci spanning 20 chromosomal regions, enriching our understanding of the genetic landscape under scrutiny. Seven of the 49 loci identified were novel and had not been previously associated with MAFLD or CVDs. Among the 22 loci (44.9%), the selected effect alleles at the top SNPs within or near loci demonstrated effect directions that were consistent with each other which means they may concurrently increase ADs and CVDs or diminish the risk of developing ADs and CVDs. Notably, three loci, 22q13.31, 16q12.2, and 8q24.13, were associated with more than half of the trait pairs analyzed. The chromosomal region 22q13.31, in particular, is implicated in five trait pairs, excluding MAFLD-Stroke, highlighting its significance in the pleiotropic landscape. For example, *SAMM50* located at 22q13.31, studies have shown that *SAMM50* gene polymorphisms are associated with the occurrence and severity of fatty liver disease, which may be related to reduced fatty acid oxidation caused by *SAMM50* deficiency^40^. Although there is no direct evidence linking *SAMM50* to cardiovascular disease, its potential effects on heart development and function by regulating mitochondrial autophagy may influence cardiovascular disease risk^41^. Moreover, the *FTO* gene, located in the 16q12.2 region, is another gene of interest due to its impact on energy intake and expenditure, potentially increasing susceptibility to both MAFLD and CVDs^42,43^. Moreover, the *TRIB1* gene, located at 8q24.13, plays a critical role in regulating lipid metabolism. It influences downstream signaling pathways such as PI3K-AKT and MAPK, which promote the accumulation of liver lipids and increase the risk of MAFLD^44,45^. Additionally, polymorphisms in the *TRIB1* gene have been linked to an increased risk of CAD and Stroke. Specifically, the A allele at rs2954029 has been significantly associated with these conditions; individuals carrying the A allele exhibit a higher risk of both CAD and stroke^46^.

Functional annotation via FUMA identified 24 intronic SNPs, 14 intergenic SNPs, and 5 exonic SNPs. For example, the 22q13.31 region includes the exonic SNP rs1007863, which showed significant eQTL associations in several tissues: adipose subcutaneous (*P* = 2.38×10^-6^), adipose visceral omentum (*P* = 2.16×10^-15^), whole blood (*P* = 3.57×10^-5^), artery aorta (*P* = 3.57×10^-5^), and artery tibial (*P* = 1.42×10^-12^). Notably, the P-value for adipose visceral omentum was used as the P-value for this region in MAFLD-AF for PLACO results, indicating that this P-value is considered the most appropriate or representative indicator, suggesting that visceral tissues are importantly associated with this trait pair. Seven of all pleiotropic SNPs were identified as potentially deleterious, with a CADD score greater than 12.37. Notably, rs28929474 exhibited the highest CADD score of 20.2, suggesting its potential association with the development of MAFLD and CAD and its deleterious effects. Furthermore, based on RDB scores, two SNPs were found to potentially impact transcription factor binding, with rs10401969 showing the most significant effect (RBD = 1f). Further colocalization analysis revealed evidence of colocalization in 13 of the 49 pleiotropic loci (PPH4 > 0.7). Notably, the 16q12.2 region has demonstrated a robust colocalization signal across all four trait pairs it encompasses (with a posterior probability of heterogeneity, PPH4, ranging from 0.766 to 0.890).

### Gene-level annotation of MAFLD and CVDs

Using MAGMA, we identified 34 potential pleiotropic genes (28 of which are unique) between MAFLD and CVDs after Bonferroni correction, which is situated on or overlaps with 49 previously identified pleiotropic loci. Among these, 2 are novel unique genes, 3 have newly unique been associated with CVDs, and 23 are newly unique implicated in MAFLD. Notably, three genes—*SAMM50*, PNPLA3, and *FTO*—were detected in two or more trait pairs. Further, of the genes identified by MAGMA, 32 were corroborated by FUMA’s locus mapping. The pervasive impact of *SAMM50* across multiple traits underscores its significance and makes it a primary focus of our ongoing investigation.

We performed tissue-specific analyses employing LDSC-SEG to identify the tissues associated with each disease. After adjusting for multiple comparisons using the FDR method, significant associations were observed solely with CVDs. Specifically, for AF, the auricle and the left ventricle were identified as the primary tissues driving the condition, suggesting that functional changes in these areas may play a crucial role in its development and maintenance. Similarly, in CAD, the predominant tissues associated with the disease were located in arterial regions, including the aorta, coronary artery, and anterior tibial artery, with the anterior tibial artery demonstrating the most significant impact. Unfortunately, while MAFLD showed enrichment in the anterior cingulate cortex, cerebral cortex, and frontal cortex, these did not meet the significant threshold for correction. In all, afte integrating the results of tissue enrichment analysis with tissues commonly implicated in clinical practice for these conditions, we finally selected 10 tissues for subsequent analysis.

Given the limitations of MAGMA, which assigns SNPs based solely on proximity to genes and may overlook the effects of distant gene regulation, we employed e-MAGMA to overcome these deficiencies. By integrating eQTL data from 10 distinct tissues, e-MAGMA enabled us to identify 161 tissue-specific genes, including 59 that were unique. Notably, *IRAK1BP1*, *ATP13A1*, *SAMM50* and *NME7* were identified in more than half of the trait pairs. *IRAK1BP1* exhibits high specificity to nine tissues except EBV-transformed lymphocytes and has not previously been associated with MAFLD or any CVDs. However, its involvement in inflammation suggests a potential role in these diseases. Our findings were further validated using FUMA’s GTExV8 data through e-MAGMA, confirming the identification of 78 tissue-specific genes, including *SAMM50* at 22q13.31. Subsequent TWAS analysis of the original GWAS results was used to validate the EMAGMA analysis, and 300 tissue-specific genes were identified as new genes for MAFLD and 185 as new genes for CVDs.

Finally, 17 pleiotropic genes (14 unique) were identified by MAGMA and e-MAGMA. *SAMM50* at 22q13.31 was the only gene that appeared in multiple trait pairs (except for MAFLD-PAD and MAFLD-Stroke), while other genes appeared only in single trait pairs.

### Functional enrichment analysis of pleiotropic genes in MAFLD and CVDs

Understanding how identified genes collectively fulfill specific biological functions through shared pathways enhances our ability to derive meaningful interpretations from genomic data. The tool ToppFun assists in mapping these genes to their respective enrichment pathways and biological processes, which can reveal the activity levels or enrichment of certain pathways. Notably, three pathways were significantly enriched: membrane organization, endoplasmic reticulum tubular network organization, and the establishment of protein localization to the membrane. Protein localization is critical in the accumulation of lipid droplets near mitochondria; it helps protect mitochondrial function by sequestering toxic lipids. This protective mechanism is crucial for managing lipid accumulation in the liver and other tissues, where excessive lipid buildup can lead to MAFLD and CVDs^47,48^. Proper protein localization, therefore, plays a pivotal role in mitigating lipid toxicity and maintaining cellular balance, thereby reducing the risk of both MAFLD and CVD.

## Discussion

This genome-wide pleiotropic association study offers compelling evidence of a shared genetic architecture and mechanisms between MAFLD and six major CVDs. We observed moderate to strong genome-wide genetic associations between MAFLD and four CVDs—CAD, HF, PAD, and VTE—with notable genetic overlap in all but the MAFLD-AF trait pair. MR analysis provided evidence of causal relationships for two trait pairs, underscoring the presence of vertical pleiotropy. Additionally, at the SNP level, our cross-trait analysis pinpointed 49 pleiotropic loci, 13 demonstrating strong evidence of colocalization. Further analysis at the gene level through MAGMA and e-MAGMA identified 45 (34 unique) significant position-specific pleiotropic genes and 161 (59 unique)tissue-specific pleiotropic genes, including 17 overlapping pleiotropic genes (14 unique because *SAMM50* was identified in 4 trait pairs). Moreover, at the biological pathway level, protein localization to membrane signaling pathways was identified as potentially pivotal in the co-pathogenesis of MAFLD and CVD. These insights not only deepen our understanding of their intertwined genetic etiology but also illuminate potential targets for therapeutic intervention and preventive strategies.

In our study, bivariate LDSC revealed extensive positive genetic correlations between MAFLD and six major CVDs, among which only four trait pairs (including MAFLD-CAD, MAFLD-HF, MAFLD-PAD, and MAFLD-VTE) passed the Bonferroni correction. Further analysis using the GPA method demonstrated significant genetic overlap for all pairs except MAFLD with AF, underscoring the shared genetic underpinnings of these conditions. Interestingly, although no significant genome-wide genetic correlation was detected between MAFLD and stroke, substantial genetic overlap suggested the presence of complex genomic interactions, which was also confirmed by local genetic correlations of LAVA. In the MAFLD-stroke pair, we identified loci with mixed effect directions—two positively and three negatively correlated—suggesting that local mixed effects might underlie the non-significant genome-wide genetic correlation, potentially driven by a range of pleiotropic variants^11^. Additionally, despite the lack of a significant association between MAFLD and AF in both LDSC and GPA analyses, our bivariate LAVA under a loose threshold indicated a positive regional genetic correlation between the two. Overall, our findings confirm that MAFLD and CVDs share substantial heritability and genetic components, elucidating the genetic basis of their high comorbidity.

The shared genetic basis between MAFLD and CVD is primarily driven by horizontal and vertical pleiotropy (i.e., causal relationships). We employed the LHC-MR method to investigate potential causal links between MAFLD and 6 major CVDs. Our analysis revealed that genetically predicted MAFLD is causally associated with an increased risk of AF and PAD. These findings are consistent with prior epidemiological evidence. A meta-analysis confirmed an association between MAFLD and an increased risk of AF^49^. Concurrently, a separate cohort study observed a high prevalence of PAD among MAFLD patients over 40 years old in the United States during a 13-year follow-up^50^. Conversely, we found no established causal relationship between MAFLD and HF or Stroke, nor was a reverse causal relationship between MAFLD and CVD supported. This contrasts with earlier MR analyses that reported a positive causal relationship between MAFLD and both HF^15^ and CAD^14^, which suggested a 15% increase in CAD incidence per unit increase in MAFLD. These discrepancies may arise from limitations in previous studies, such as potential genetic confounders and smaller sample sizes, which could impact the reliability of the exposure-outcome relationship. LHC-MR analysis is particularly effective at generating potential causal effects by excluding confounding factors, even in the face of unmeasured confounding. Leveraging extensive sample data and advanced methodologies, our study provided more robust and reliable causal effect estimates. Our study only provides partial evidence for vertical pleiotropy, suggesting that the shared genetic basis between MAFLD and CVD is primarily driven by horizontal pleiotropy.

Horizontal pleiotropy analysis further elucidated the common genetic architecture between MAFLD and CVD by discovering pleiotropic variants and loci, pleiotropic genes, and biological pathways between the two. Further investigation through horizontal pleiotropy analysis has confirmed their extensive comorbidity, identifying widespread pleiotropic variants across several key loci, such as 22q13.31, 16q12.2, and 8q24.13, which are prominently associated with more than half of the trait pairs analyzed. Notably, the locus at 16q12.2—associated with all trait pairs except MAFLD-CAD and MAFLD-Stroke—maps to the *FTO* gene. This gene plays a critical role in lipid metabolism by enhancing oxidative stress and increasing lipogenesis in hepatocytes, thereby exacerbating MAFLD progression^51^. Moreover, according to a meta-analysis of 10 observational studies (19,153 CVD cases and 103,720 controls), variants in the *FTO* gene have been shown to significantly elevate CVD risk, independent of BMI and other conventional risk factors^52^. Moreover, in a large prospective longitudinal study of Type 2 diabetes (T2D) patients, variants in the *FTO* gene were also found to increase the risk of myocardial infarction and cardiovascular death by influencing a dyslipidemic phenotype typical of insulin resistance^53^. The above evidence suggests that the lipid metabolism process can be significantly changed by affecting the expression of the *FTO* gene, thereby improving MAFLD and reducing the risk of cardiovascular death.

In our gene-level analysis using MAGMA and e-MAGMA, we identified significant pleiotropic genes at the 22q13.31 locus, including patatin-like phospholipase-domain containing protein 3 (*PNPLA3*) and sorting and assembly machinery component 50 (*SAMM50*). These genes play crucial roles in the comorbidity between MAFLD and CVDs. *PNPLA3*, which exhibits weak hydrolytic activity towards glycerolipids, has been linked to the development of MAFLD and CVDs, excluding PAD and Stroke. This gene is associated with a missense variant closely linked to triacylglycerol (TAG) accumulation in the liver—a major genetic risk factor for steatotic liver diseases^54,55^. *PNPLA3* competitively displaces adipose triglyceride lipase (ATGL) from lipid droplets, reducing lipolytic activity and promoting the progression of MAFLD^54^. Intriguingly, a GWAS meta-analysis revealed that the *PNPLA3* G-allele rs738409 offers modest protection against CAD by reducing triglyceride breakdown in the liver, thus affecting the secretion of very low-density lipoprotein particles and the development of atherosclerosis^56^. Additionally, our e-MAGMA analysis identified *SAMM50* at this locus as influencing most trait pairs. As an essential component of the Sorting and Assembly Machinery (SAM) complex on the outer mitochondrial membrane, *SAMM50* is vital for β-barrel protein biogenesis and interacts directly with the translocator of the outer mitochondrial membrane (TOM) complex^41^. It plays a significant role in fatty acid β-oxidation and, when deficient, can lead to intracellular lipid accumulation, exacerbating MAFLD^40^. Moreover, *SAMM50*’s involvement in mitochondrial dysfunction may impair the removal of reactive oxygen species (ROS), further contributing to the progression of MAFLD^57^ and increasing susceptibility to HF by impairing mitochondrial function^41^. This mitochondrial pathway regulated by *SAMM50* offers a theoretical basis for the observed comorbidity between MAFLD and HF in our study, underscoring the potential for targeted therapies at this genetic locus.

In our pathway-level analysis, we discovered a crucial role for the gene *SAMM50* in mediating protein localization to membranes, a process integral to the pathogenesis of MAFLD. This protein localization is especially important in how lipid droplets accumulate near mitochondria, which may protect mitochondrial function by sequestering toxic lipid species. Proteins that migrate to these lipid droplets at membrane contact sites are prime candidates for facilitating this protective mechanism^47,48^. In the liver, an excessive influx of lipids leads to the accumulation of triglyceride (TG)-rich lipid droplets, a hallmark of MAFLD^47^. Variations in genes associated with lipid droplet formation, such as PNPLA3, acting as both phospholipase and acyl transfer enzyme, is an important regulating factor for TG. It limits the activity of triglyceride hydrolases, leading to TG accumulation in the liver, thus predisposing to MAFLD^58^. Moreover, when lipid droplets amass in peripheral organs, the overflow of toxic lipids can contribute to CVDs. ATGL plays a pivotal role in this context by hydrolyzing TG within these droplets. It activates the PPARα/peroxisome proliferator-activated receptor-γ coactivator 1 (*PGC1*) complex in cardiomyocytes. A deficiency in cardiac ATGL leads to reduced *PGC1* expression, resulting in mitochondrial dysfunction and lipid accumulation, which can progress to HF^59^. In conclusion, the localization of contact site proteins related to lipid droplets is instrumental in preventing lipid overflow and safeguarding mitochondrial function. This mechanism significantly reduces the risk of both MAFLD and CVD by maintaining crucial cellular balances and interactions.

Our study has several limitations. First, our study population was confined to individuals of European ancestry, necessitating further research across other ethnic groups to determine the universal applicability of our findings. Second, the datasets used for MAFLD and CVD included some cases with existing or potential co-morbid conditions, which may have biased our investigation of genetic overlap. Third, our analysis was limited to common genetic variants, highlighting the need for future studies to explore the impact of rare variants, which may also significantly influence the high comorbidity observed between MAFLD and CVD. Finally, while we reported some statistically significant findings, further investigation is required to assess their clinical relevance and implications.

In summary, our study demonstrates extensive genetic overlap, partial genetic correlation, and causal relationships between MAFLD and six major CVDs. Identification of the pleiotropic gene, *SAMM50*, at pleiotropic risk loci 22q13.31, shared between MAFLD and CVD, suggesting a common biological mechanism, namely the establishment of protein localization to the membrane, uncovering indications of shared genetic foundation for MAFLD and CVDs. This research enhances the shared genetic map of these diseases and offers new perspectives on preventing and treating their co-morbidities.

## Data Availability

All used data in the study are available to be download online. The datasets used in this study are publicly available from the following sources: atrial fibrillation data can be accessed at the GWAS Catalog (GCST006414) through https://www.ebi.ac.uk/gwas/studies/GCST006414; coronary artery disease data are available at https://cvd.hugeamp.org/datasets.html; venous thromboembolism data are accessible at https://www.decode.com/summarydata/; heart failure data can be found at the GWAS Catalog (GCST009541) via https://www.ebi.ac.uk/gwas/studies/GCST009541; peripheral artery disease data are provided at https://cvd.hugeamp.org/datasets.html; stroke data can be obtained at https://www.ebi.ac.uk/gwas/studies/GCST90104539; and metabolic dysfunction-associated fatty liver disease data are available at https://www.ebi.ac.uk/gwas/publications/34841290.

https://cvd.hugeamp.org/datasets.html

https://www.decode.com/summarydata/

https://www.ebi.ac.uk/gwas/studies/GCST009541

https://cvd.hugeamp.org/datasets.html

https://www.ebi.ac.uk/gwas/studies/GCST90104539

https://www.ebi.ac.uk/gwas/publications/34841290

## Abbreviations

AF: atrial fibrillation
ApoC3: apolipoprotein C3
ATGL: adipose triglyceride lipase
CVDs: cardiovascular diseases
CAD: coronary artery disease
CRP: C-Reactive Protein
CADD: Combined Annotation-Dependent Depletion
e-MAGMA: eQTL-informed MAGMA
eMERGE: Electronic Medical Records and Genomics
EstBB: Estonian Biobank
FUMA: Functional Mapping and Annotation
FDR: false discovery rate
GPA: Genetic Analysis incorporating Pleiotropy and Annotation
GWAS: genome-wide association studies
*h*^2^: SNP-based heritability
HF: heart failure
HUNT: Health Study Nord-Trøndelag
HERMES: Heart Failure Molecular Epidemiology for Therapeutic Targets
IVW: Inverse Variance Weighting
IL-1: interleukin-1
LDSC: linkage disequilibrium score regression
LD: linkage disequilibrium
LDSC-SEG: LDSC applied to specifically expressed genes
LRT: likelihood ratio test
LAVA: Local Analysis of [co]Variant Annotation
LHC-MR: Latent Heritable Confounder Mendelian randomization
MAFLD: metabolic dysfunction-associated fatty liver disease
MR: Mendelian Randomization
MGI: Michigan Genome Initiative
MAF: minor allele frequency
MHC: major histocompatibility complex
MAGMA: Multi-marker Analysis of GenoMic Annotation
NAFLD: Non-alcoholic fatty liver disease
OR: odds ratios
PAD: peripheral artery disease
PLACO: Pleiotropic Analysis under a Composite Null Hypothesis
RDB: RegulomeDB
PP: posterior probabilities
*PNPLA3*: patatin-like phospholipase-domain containing protein 3
*PGC1*: PPARα/peroxisome proliferator-activated receptor-γ coactivator 1
ROS: reactive oxygen species
SNPs: single nucleotide polymorphisms
*SAMM50*: sorting and assembly machinery component 50
SAM: Sorting and Assembly Machinery
TWAS: transcriptome-wide association study
ToppFun: ToppGene functional annotation tool
TFBS: transcription factor binding sites
T2D: Type 2 diabetes
TAG: triacylglycerol
TOM: translocator of the outer mitochondrial membrane
TG: triglyceride
UKB: UK Biobank
VTE: venous thromboembolism)

## Acknowledgements

We would like to thank all GWAS authors who provided summary statistics for this study. We thank the Electronic Medical Records and Genomics (eMERGE) Network, the UK Biobank (UKB), the Estonian Biobank (EstBB), and the FinnGen Consortium for providing us with data on MAFLD, without whom this work would not have been possible. We would also like to thank the Health Study Nord-Trøndelag (HUNT), deCODE, Michigan Genome Initiative (MGI), DiscovEHR, UKB, and AFGen consortium for providing AF data; the CARDIoGRAMplusC4D and UKB consortium for providing CAD data; the deCODE for providing VTE data; the Heart Failure Molecular Epidemiology for Therapeutic Targets (HERMES) consortium for providing HF data; the Cardiovascular Disease Knowledge Portal (CVDKP) website for providing PAD data; and the GIGASTROKE consortium for providing stroke data.

---

**Table 1.** Genetic Correlation and Genetic Overlap Estimations Between 6.

| Trait pair | Genetic correlation |  |  |  | Genetic overlap |  |  |
| --- | --- | --- | --- | --- | --- | --- | --- |
|  | Genetic correlation (SE) | *P* value for LDSC | Intercept (SE) | *P* value for intercept | PM 11 | PAR<sup>b</sup> | *P* value for GPA |
| MAFLD-AF | 0.1381<br>(0.0891) | 0.121 | 0.0080<br>(0.0053) | 0.131 | 0.0000 | 0.0004 | 9.55×10<sup>-2</sup> |
| MAFLD-CAD<sup>c,d</sup> | 0.6266<br>(0.1375) | 5.18×10<sup>-6</sup> | 0.0018<br>(0.0059) | 0.760 | 0.0036 | 0.0424 | 7.12×10<sup>-31</sup> |
| MAFLD-HF<sup>c,d</sup> | 0.5725<br>(0.1576) | 3.00×10<sup>-4</sup> | 0.0058<br>(0.0045) | 0.197 | 0.0001 | 0.0010 | 1.12×10<sup>-11</sup> |
| MAFLD-PAD<sup>c,d</sup> | 0.6259<br>(0.2099) | 2.90×10<sup>-3</sup> | 0.0075<br>(0.0053) | 0.157 | 0.0001 | 0.0029 | 1.32×10<sup>-03</sup> |
| MAFLD-Stroke<sup>d</sup> | 0.2783<br>(0.1353) | 3.98×10<sup>-2</sup> | 0.0057<br>(0.0045) | 0.205 | 0.0001 | 0.0007 | 6.96×10<sup>-6</sup> |
| MAFLD-VTE<sup>c,d</sup> | 0.4844<br>(0.1220) | 7.19×10<sup>-5</sup> | 0.0025<br>(0.0051) | 0.624 | 0.0002 | 0.0025 | 4.66×10<sup>-10</sup> |

Note:
a. Genetic correlation and genetic overlap were estimated by LDSC and GPA methods, respectively. Bonferroni-corrected significance threshold was set at $P \leq 8.33 \times 10^{-3}$ (0.05/6), producing a final union set of 5 pairwise traits with significant genetic correlation or genetic overlap for subsequent analysis.
b. We introduced PAR as $PM_{11} / (PM_{10} + PM_{01} + PM_{11})$ to represent the proportion of pleiotropic single-nucleotide variations (SNPs) associated with both traits against the proportion of SNPs associated with at least 1 trait.
c. Pairwise trait with significant genetic correlation.
d. Pairwise trait with significant genetic overlap.
Abbreviations: MAFLD, Metabolic dysfunction-Associated fatty liver disease; AF, Atrial fibrillation; CAD, Coronary artery disease; HF, Heart failure; PAD, Peripheral artery disease; VTE, Venous thromboembolism; LDSC, linkage disequilibrium score regression; GPA, genetic analysis incorporating pleiotropy and annotation method; PAR, pleiotropy association ratio; PM 11, proportion of genetic variants associated with both traits.

**Figure 1.**
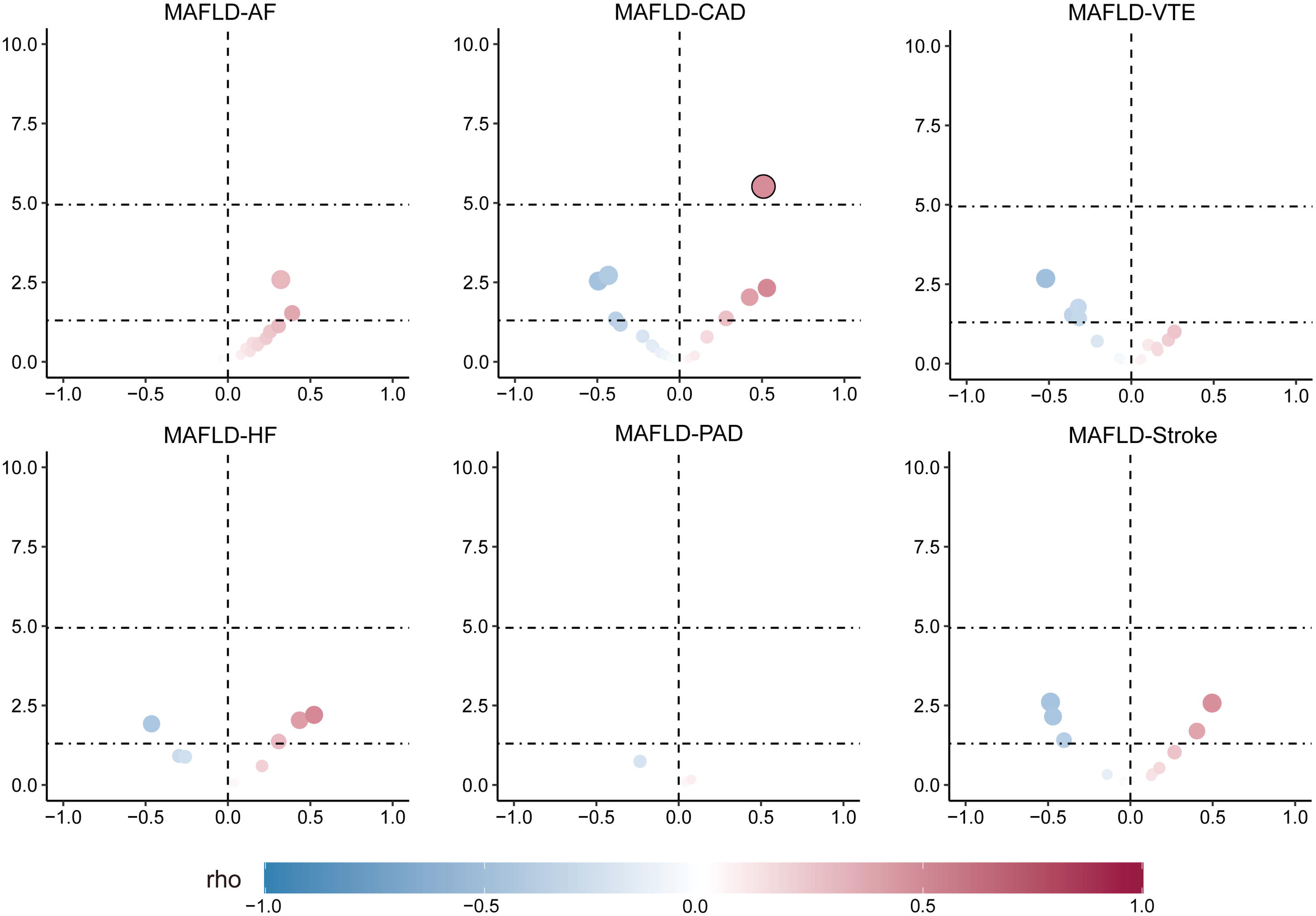
Metabolic dysfunction-associated fatty liver disease and six major cardiovascular diseases estimated by local genetic correlation. LAVA volcano plots showing local genetic correlation coefficients (local-r_g_s, y-axis) for MAFLD and CVDs with -log10 p-values for each trait pair of analyses for each locus. Loci above the horizontal line are significant at P < 0.05 (negative correlation for blue dots, positive correlation for red dots). Larger points with black circles indicate loci significantly associated after Bonferroni correction (P = 6.49×10^-4^ = 0.05 / 77). LAVA-estimated local-rgs is shown on the blue-red scale. MAFLD, metabolic dysfunction-associated fatty liver disease; AF, atrial fibrillation; CAD, coronary artery disease; VTE, venous thromboembolism; HF, heart failure; PAD, peripheral arterial disease.

**Figure 2.**
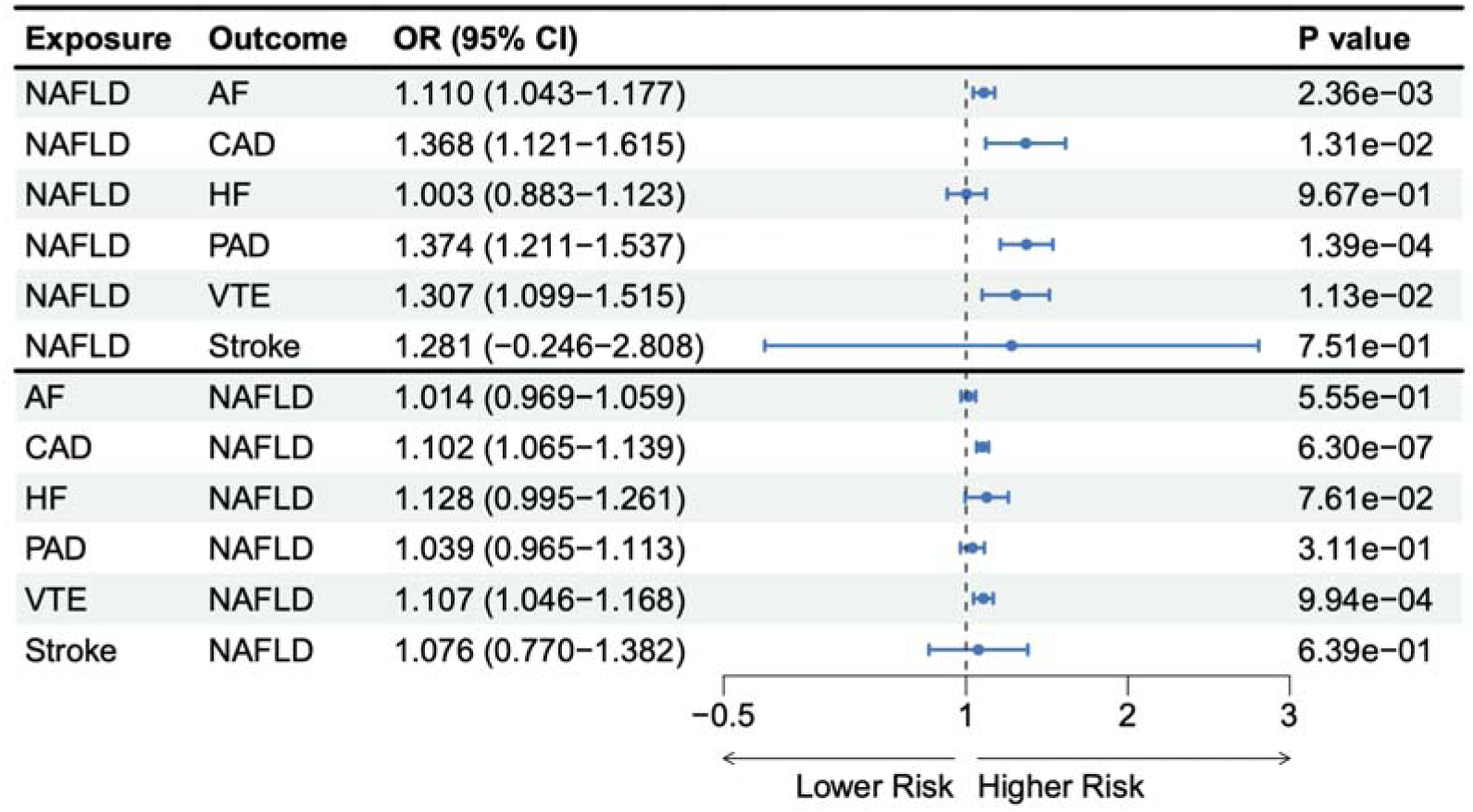
Inference of bidirectional causal relationship between metabolic dysfunction-associated fatty liver disease and six major cardiovascular diseases. Forest plots of the causal relationship between MAFLD and six major CVDs using the LHC-MR method. The estimates presented in the forest plot were obtained using the LHC-MR method. A positive association is indicated by the odd ratio (OR > 1), while a negative association is indicated by OR < 1. The results of forward causality from the LHC-MR method are in the up panel, and the results of reverse causality from the LHC-MR method are in the down panel. MAFLD, metabolic dysfunction-associated fatty liver disease; AF, atrial fibrillation; CAD, coronary artery disease; VTE, venous thromboembolism; HF, heart failure; PAD, peripheral arterial disease.

**Figure 3.**
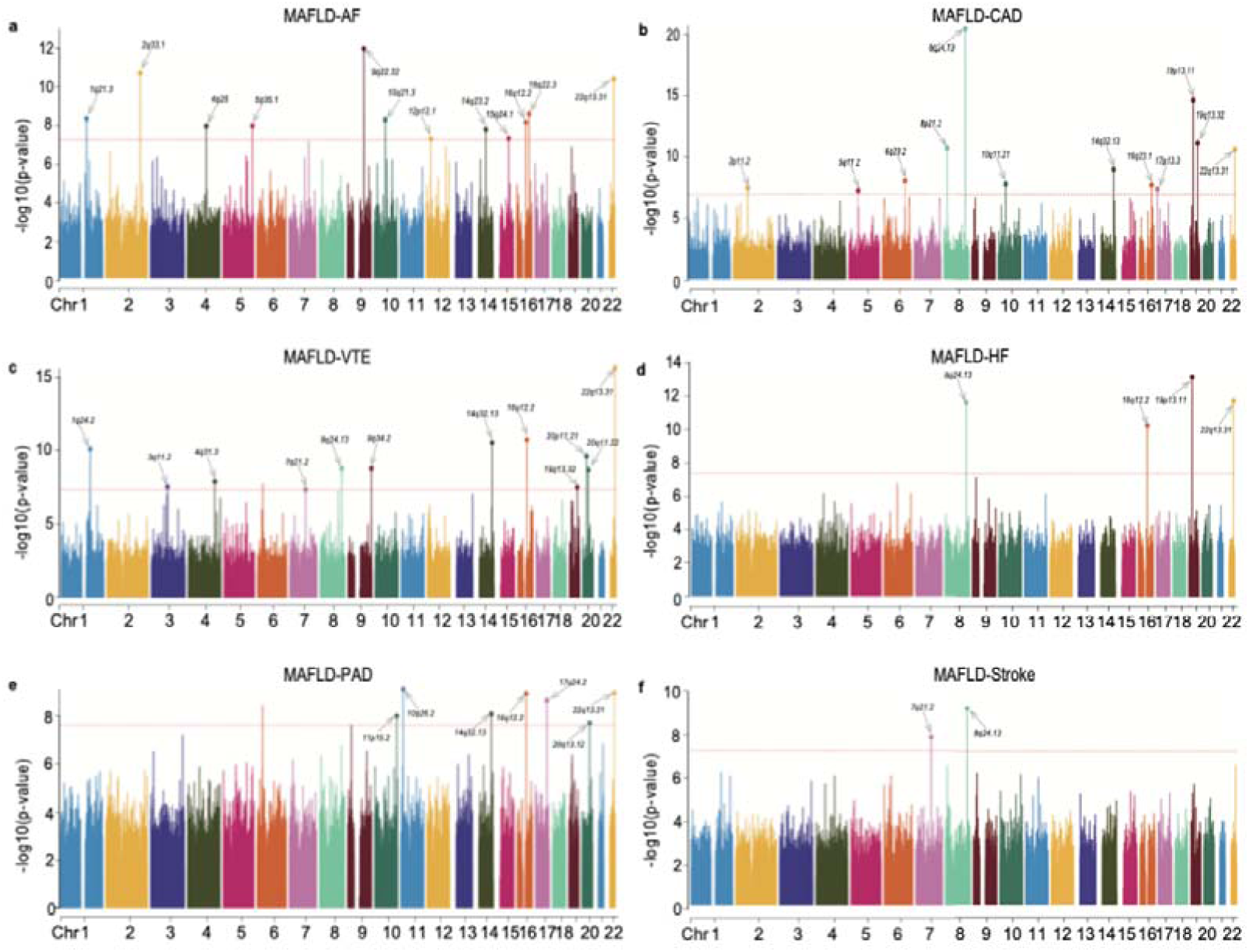
Manhattan plots for the PLACO results of metabolic dysfunction-associated fatty liver disease and six major cardiovascular diseases. Manhattan plots reflect chromosomal position (x-axis) and negative log10-transformed P-values (y-axis) for each SNP. The horizontal dashed red line indicates the genome-wide significant P-value of -log10 (5×10^-8^). The r^2^ threshold for defining independent significant SNPs was set to 0.2, and the maximum distance between LD blocks merged into one locus was set to 500 kb. The independent genome-wide significant associations with the smallest P-value (Top lead SNP) are encircled in a colorful circle. Only SNPs shared across all summary statistics were included. MAFLD, metabolic dysfunction-associated fatty liver disease; AF, atrial fibrillation; CAD, coronary artery disease; VTE, venous thromboembolism; HF, heart failure; PAD, peripheral arterial disease.

**Figure 4.**
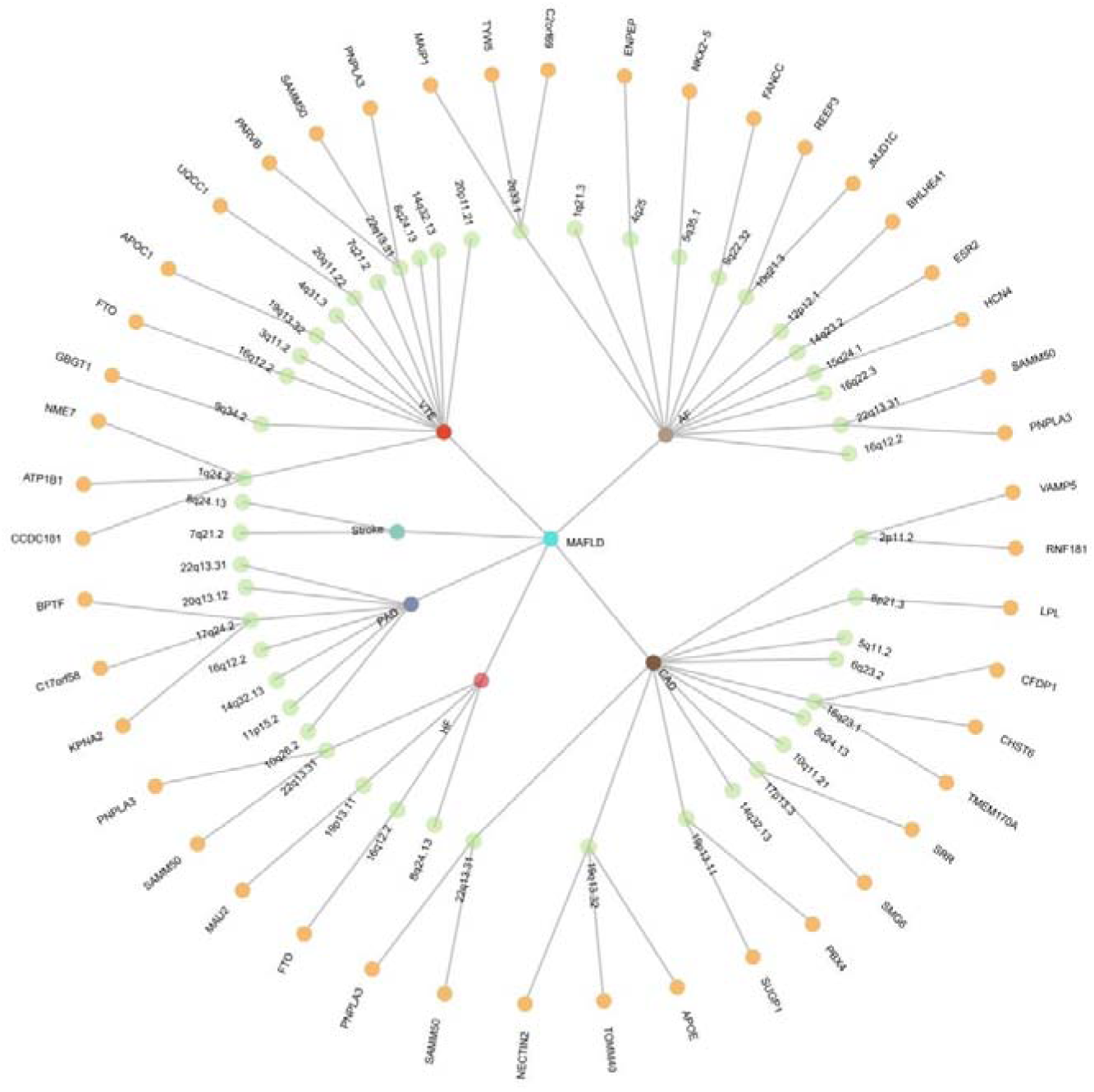
The overall situation of the pleiotropy association between metabolic dysfunction-associated fatty liver disease and six major cardiovascular diseases. A circular dendrogram showing the shared genes between MAFLD (center circle) and each of six CVDs (first circle), resulting in six pairs. A total of 49 shared loci were identified across six trait pairs (third circle), mapped to 45 nominally significant pleiotropic genes (34 were Bonferroni-corrected significant) identified by multimarker analysis of GenoMic annotation (MAGMA). For the trait pairs with more than three pleiotropic genes, we only showed the top 3 pleiotropic genes according to the prioritization of candidate pleiotropic genes (fourth circle). MAFLD, metabolic dysfunction-associated fatty liver disease; AF, Atrial fibrillation; CAD, Coronary artery disease; VTE, Venous thromboembolism; HF, Heart failure; PAD, Peripheral artery disease.

