## Supplementary Information for "Genome-wide insights into the shared genetic landscape between metabolic dysfunction-associated fatty liver disease and cardiovascular diseases"

**Supplementary Figures**

**[Supplementary Fig 1:](#_Toc181137725)** [Locus comparing plots for the shared causal variant for the associations of metabolic dysfunction-associated fatty liver disease and atrial fibrillation	2](#_Toc181137725)

**[Supplementary Fig 2:](#_Toc181137725)** [Locus comparing plots for the shared causal variant for the associations of metabolic dysfunction-associated fatty liver disease and coronary artery diseases](#_Toc181137725)	6

**[Supplementary Fig 3:](#_Toc181137725)** [Locus comparing plots for the shared causal variant for the associations of metabolic dysfunction-associated fatty liver disease and venous thromboembolism	1](#_Toc181137725)0

**[Supplementary Fig 4:](#_Toc181137725)** [Locus comparing plots for the shared causal variant for the associations of metabolic dysfunction-associated fatty liver disease and heart failure	1](#_Toc181137725)4

**[Supplementary Fig 5:](#_Toc181137725)** [Locus comparing plots for the shared causal variant for the associations of metabolic dysfunction-associated fatty liver disease and peripheral artery disease.	1](#_Toc181137725)6

**[Supplementary Fig 6:](#_Toc181137725)** [Locus comparing plots for the shared causal variant for the associations of metabolic dysfunction-associated fatty liver disease and stroke	1](#_Toc181137725)9

**Supplementary figure**

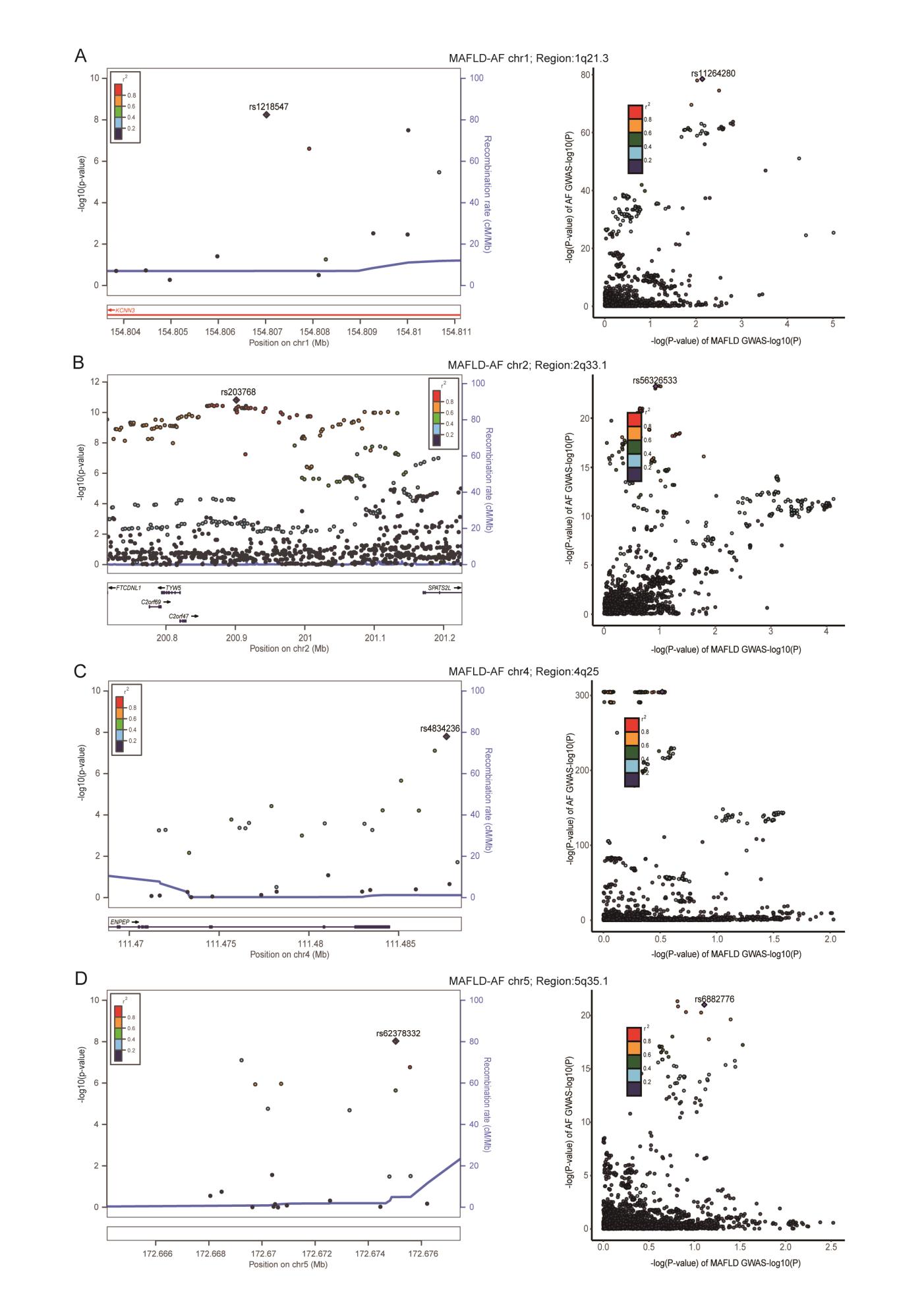

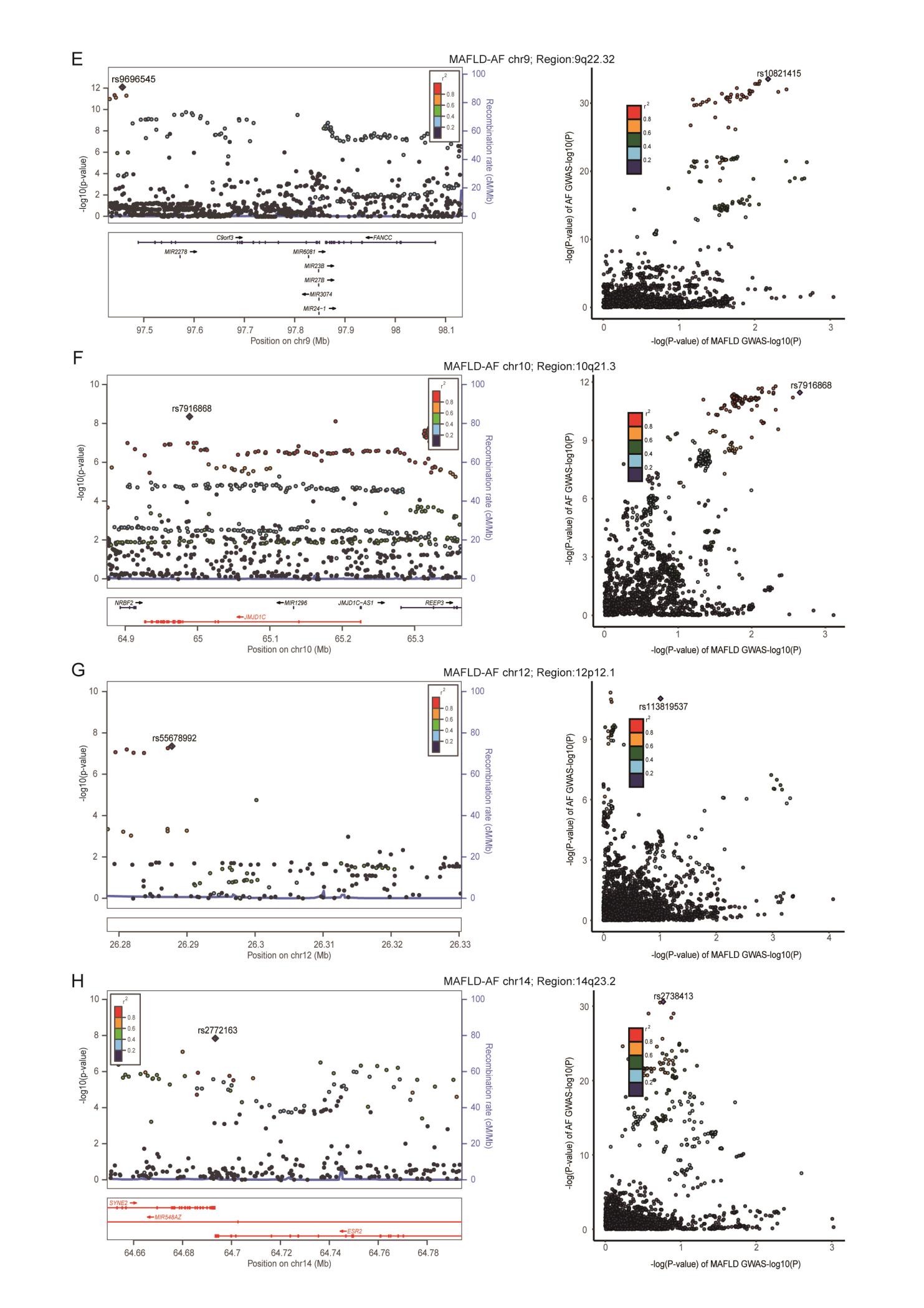

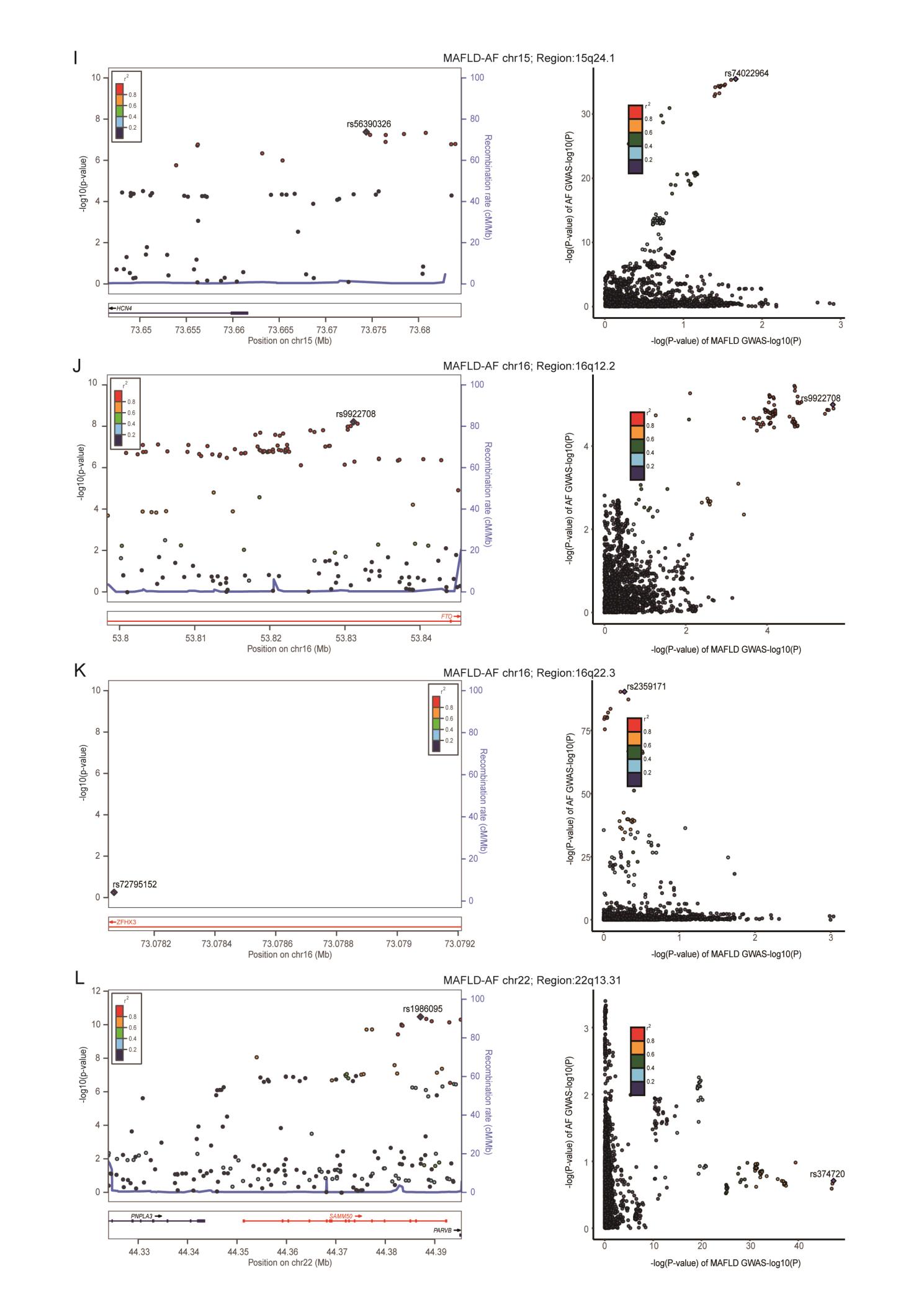

**Supplementary Fig. 1. Locus comparing plots for the shared causal variant for the associations of metabolic dysfunction-associated fatty liver disease and atrial fibrillation.**

A total of 49 genomic loci were identified to have strong evidence between MAFLD and CVDs (PP.H4 > 0.7). The left panel depicts the PLACO results using LocusZoom plots, and the right panel compares the two single-trait GWAS statistics for each variant-trait pair using LocusCompare plots. For the LocusZoom plots, the x-axis shows the genomic position of each variant, and the y-axis shows the -log10 P value from the PLCAO results. A purple diamond represents each locus's top variant with the smallest Ppraco. The color of each variant represents its LD relationship with the top variant. For the LocusCompare plots, each dot represents a variant, and the x-axis shows the -log10 PGWAS from the corresponding GWAS for MAFLD, and the y-axis shows the -log10 PGWAS from the corresponding GWAS for CVDs. Purple diamonds also represent candidate-shared causal variants identified by pairwise colocalization analysis. The color of each variant represents its LD relationship with the candidate-shared causal variant. All genomic locations are based on reference genome hg19, and the LD calculation is based on the 1000 Genomes Project of the European population. (A) 1q21.3（rs1218549）in MAFLD-AF, (B) 2q33.1（rs203768）in MAFLD-AF, (C) 4q25（rs10213638）in MAFLD-AF, (D) 5q35.1（rs62378332）in MAFLD-AF, (E) 9q22.32（rs9696545）in MAFLD-AF, (F) 10q21.3（rs7916868）in MAFLD-AF, (G) 12p12.1（rs55678992）in MAFLD-AF, (H) 14q23.2（rs2772163）in MAFLD-AF, (H) 15q24.1（rs56390326）in MAFLD-AF, (J) 16q12.2（rs9922708）in MAFLD-AF, (K) 16q22.3（rs11641701）in MAFLD-AF, (L) 22q13.31（rs1007863）in MAFLD-AF. Detailed descriptions were provided in Supplementary Table 7.

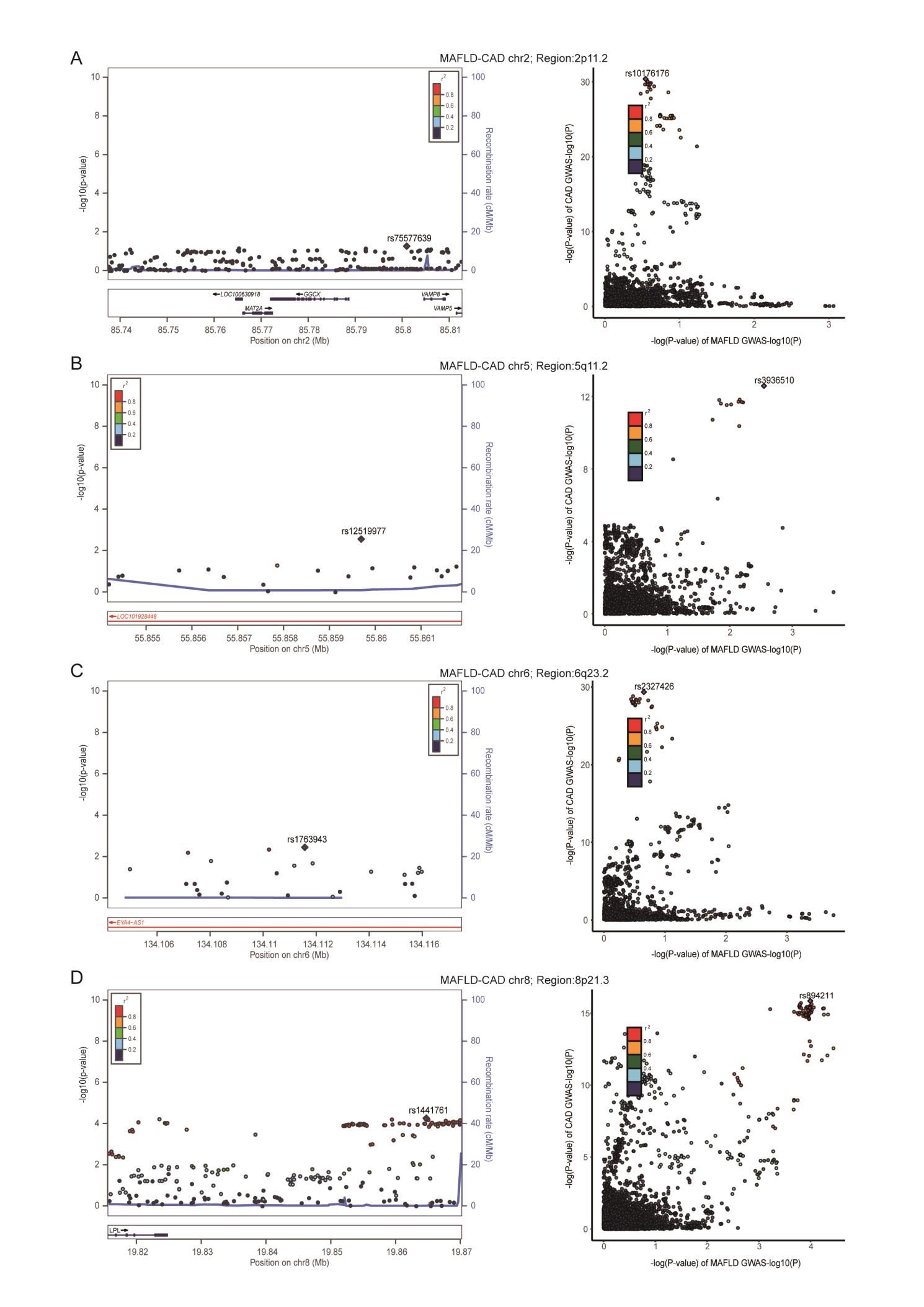

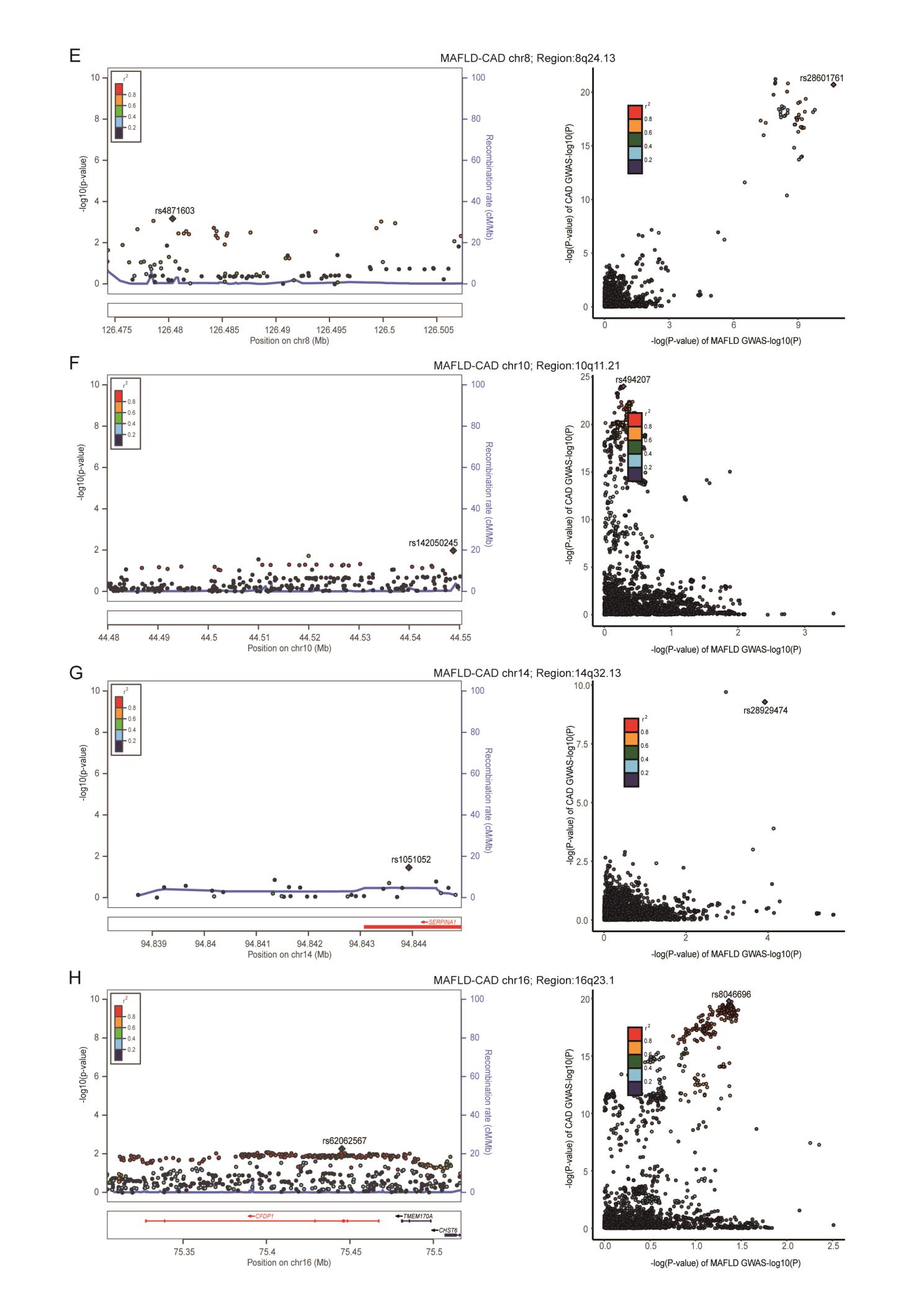

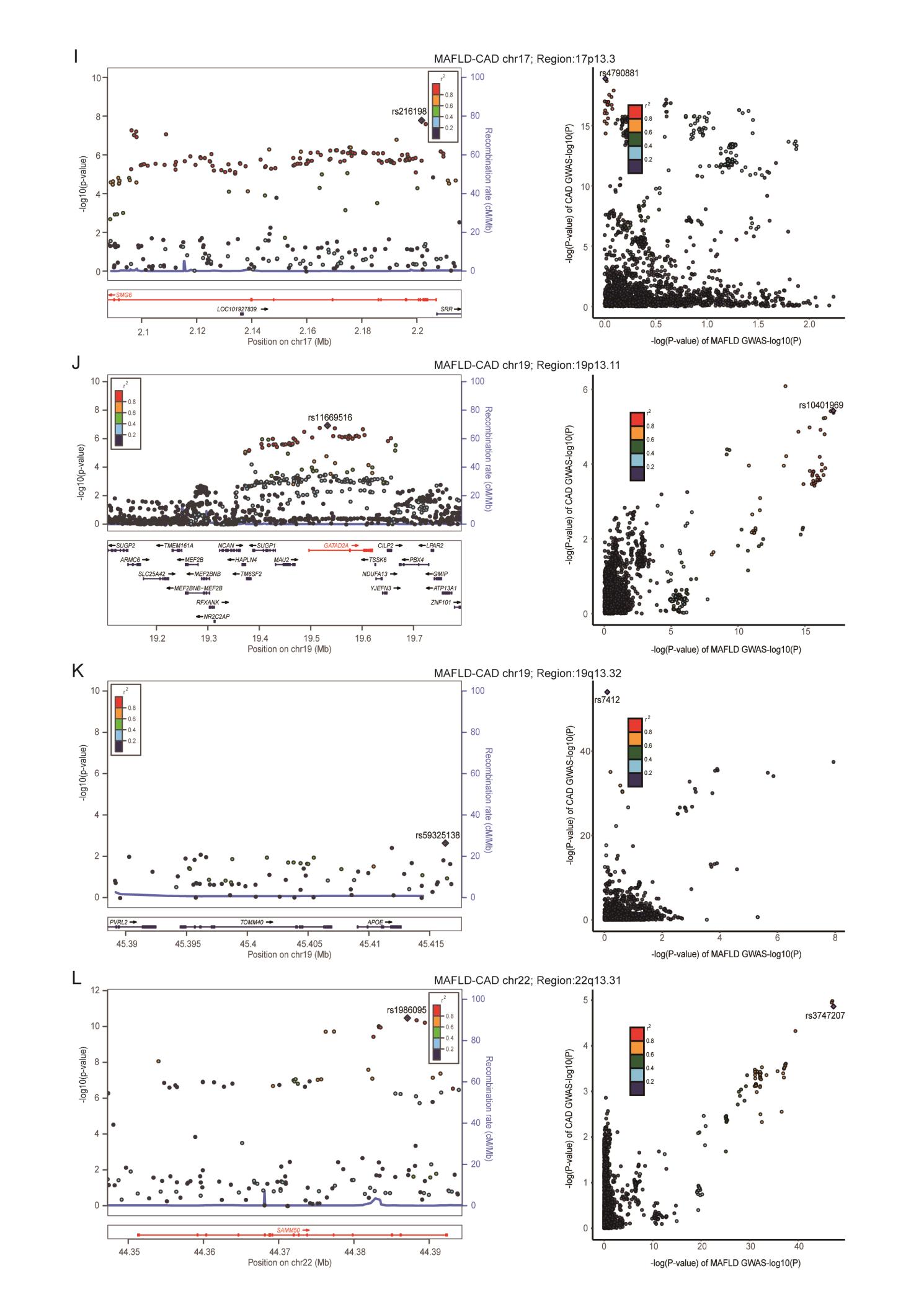

**Supplementary Fig. 2. Locus comparing plots for the shared causal variant for the associations of metabolic dysfunction-associated fatty liver disease and coronary artery disease.**

A total of 49 genomic loci were identified to have strong evidence between MAFLD and CVDs (PP.H4 > 0.7). The left panel depicts the PLACO results using LocusZoom plots, and the right panel compares the two single-trait GWAS statistics for each variant-trait pair using LocusCompare plots. For the LocusZoom plots, the x-axis shows the genomic position of each variant, and the y-axis shows the -log10 P value from the PLCAO results. A purple diamond represents each locus's top variant with the smallest Ppraco. The color of each variant represents its LD relationship with the top variant. For the LocusCompare plots, each dot represents a variant, and the x-axis shows the -log10 PGWAS from the corresponding GWAS for MAFLD, and the y-axis shows the -log10 PGWAS from the corresponding GWAS for CVDs. Purple diamonds also represent candidate-shared causal variants identified by pairwise colocalization analysis. The color of each variant represents its LD relationship with the candidate-shared causal variant. All genomic locations are based on reference genome hg19, and the LD calculation is based on the 1000 Genomes Project of the European population. (A) 2p11.2（rs62166769）in MAFLD-CAD, (B) 5q11.2（rs3936510）in MAFLD-CAD, (C) 6q23.2（rs1208256）in MAFLD-CAD, (D) 8p21.3（rs2119690）in MAFLD-CAD, (E) 8q24.13（rs28601761）in MAFLD-CAD, (F) 10q11.21（rs2818916）in MAFLD-CAD, (G) 14q32.13（rs28929474）in MAFLD-CAD, (H) 16q23.1（rs11641587）in MAFLD-CAD, (I) 17p13.3（rs216198）in MAFLD-CAD, (J) 19p13.11（rs10401969）in MAFLD-CAD, (K) 19q13.32（rs59007384）in MAFLD-CAD, (L) 22q13.31（rs2143571）in MAFLD-CAD. Detailed descriptions were provided in Supplementary Table 7.

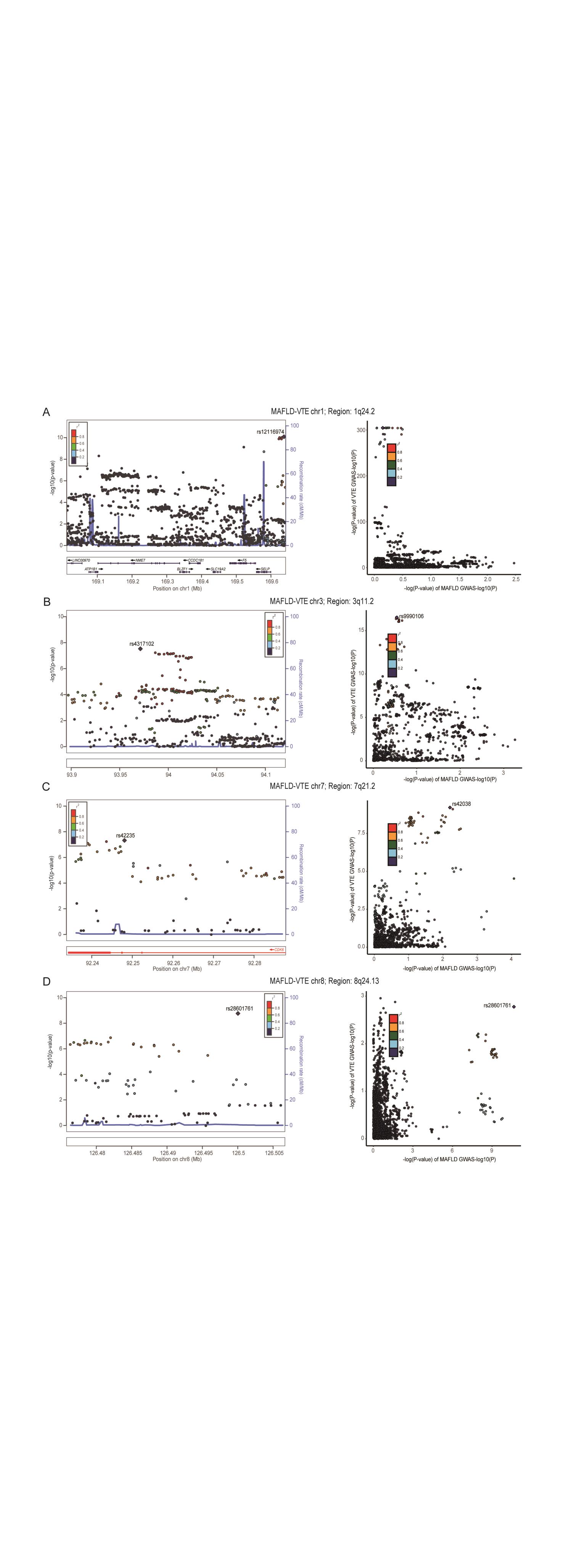

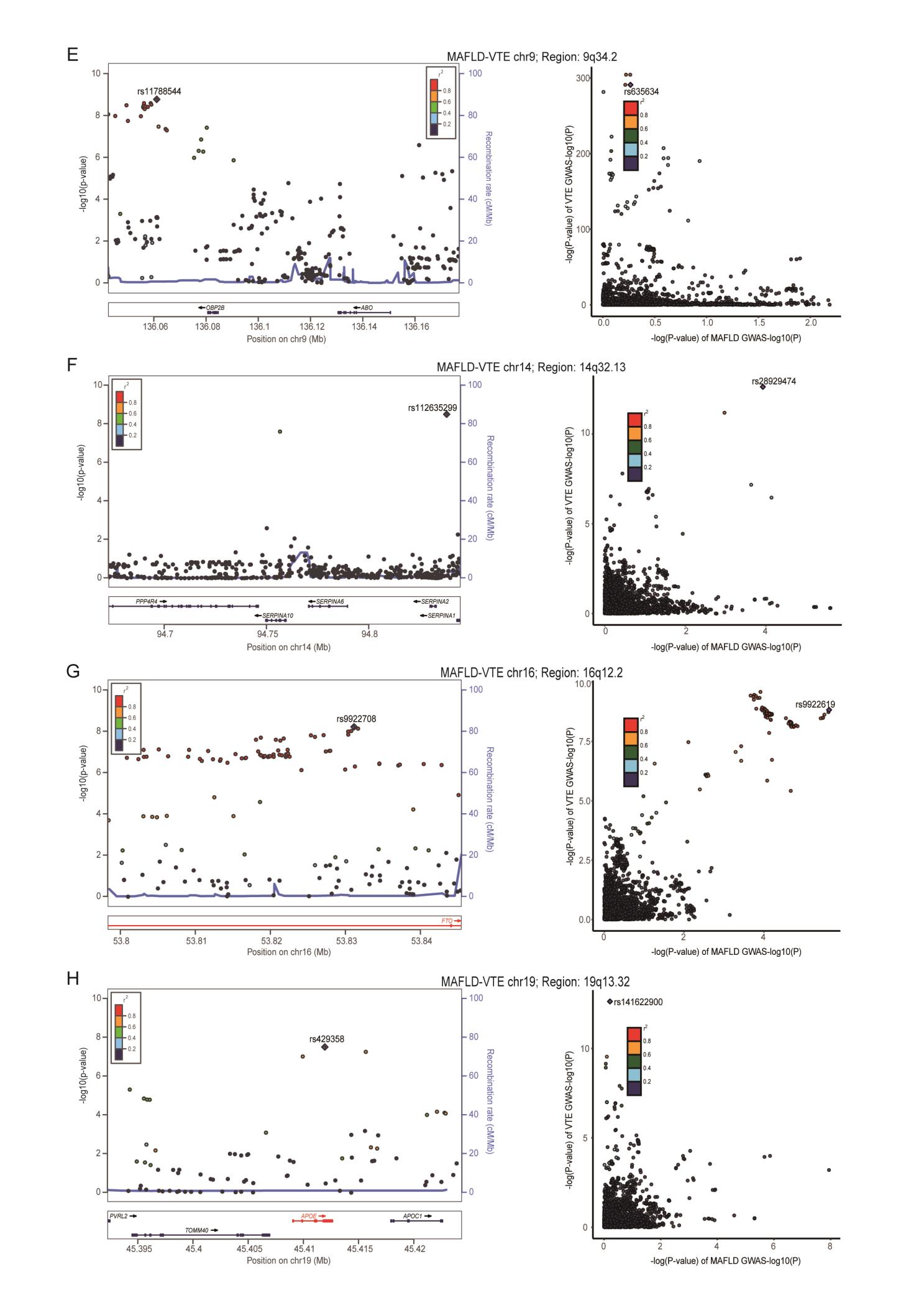

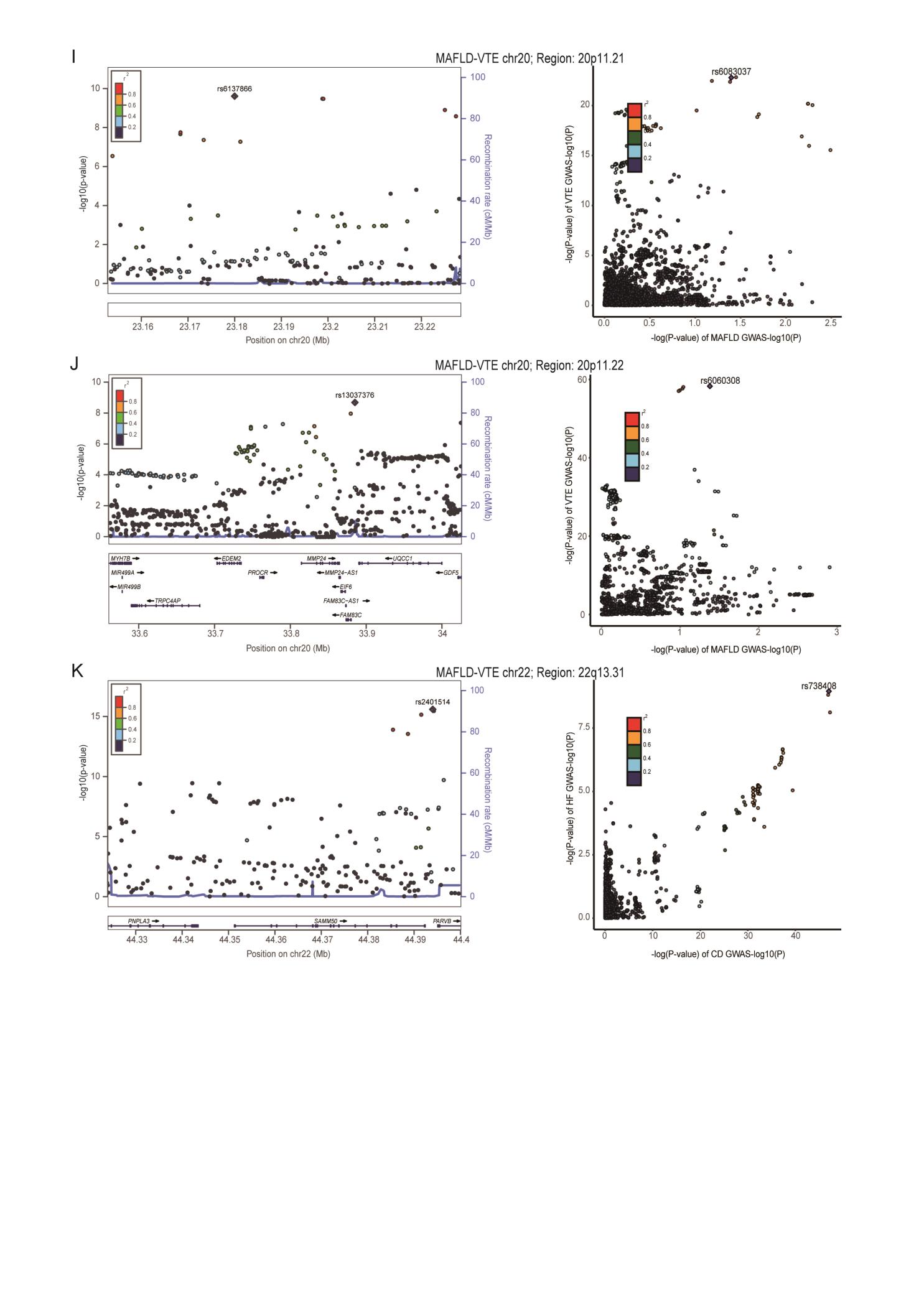

**Supplementary Fig. 3. Locus comparing plots for the shared causal variant for the associations of metabolic dysfunction-associated fatty liver disease and venous thromboembolism.**

A total of 49 genomic loci were identified to have strong evidence between MAFLD and CVDs (PP.H4 > 0.7). The left panel depicts the PLACO results using LocusZoom plots, and the right panel compares the two single-trait GWAS statistics for each variant-trait pair using LocusCompare plots. For the LocusZoom plots, the x-axis shows the genomic position of each variant, and the y-axis shows the -log10 P value from the PLCAO results. A purple diamond represents each locus's top variant with the smallest Ppraco. The color of each variant represents its LD relationship with the top variant. For the LocusCompare plots, each dot represents a variant, and the x-axis shows the -log10 PGWAS from the corresponding GWAS for MAFLD, and the y-axis shows the -log10 PGWAS from the corresponding GWAS for CVDs. Purple diamonds also represent candidate-shared causal variants identified by pairwise colocalization analysis. The color of each variant represents its LD relationship with the candidate-shared causal variant. All genomic locations are based on reference genome hg19, and the LD calculation is based on the 1000 Genomes Project of the European population. (A) 1q24.2（rs12116974）in MAFLD-VTE, (B) 3q11.2（rs4317102）in MAFLD-VTE, (C) 7q21.2（rs42235）in MAFLD-VTE, (D) 8q24.13（rs28601761）in MAFLD-VTE, (E) 9q34.2（rs11788544）in MAFLD-VTE, (F) 14q32.13（rs28929474）in MAFLD-VTE, (G) 16q12.2（rs9922619）in MAFLD-VTE, (H) 19q13.32（rs429358）in MAFLD-VTE, (I) 20p11.21（rs6137866）in MAFLD-VTE, (J) 20q11.22（rs13037376）in MAFLD-VTE, (K) 22q13.31（rs2401514）in MAFLD-VTE. Detailed descriptions were provided in Supplementary Table 7.

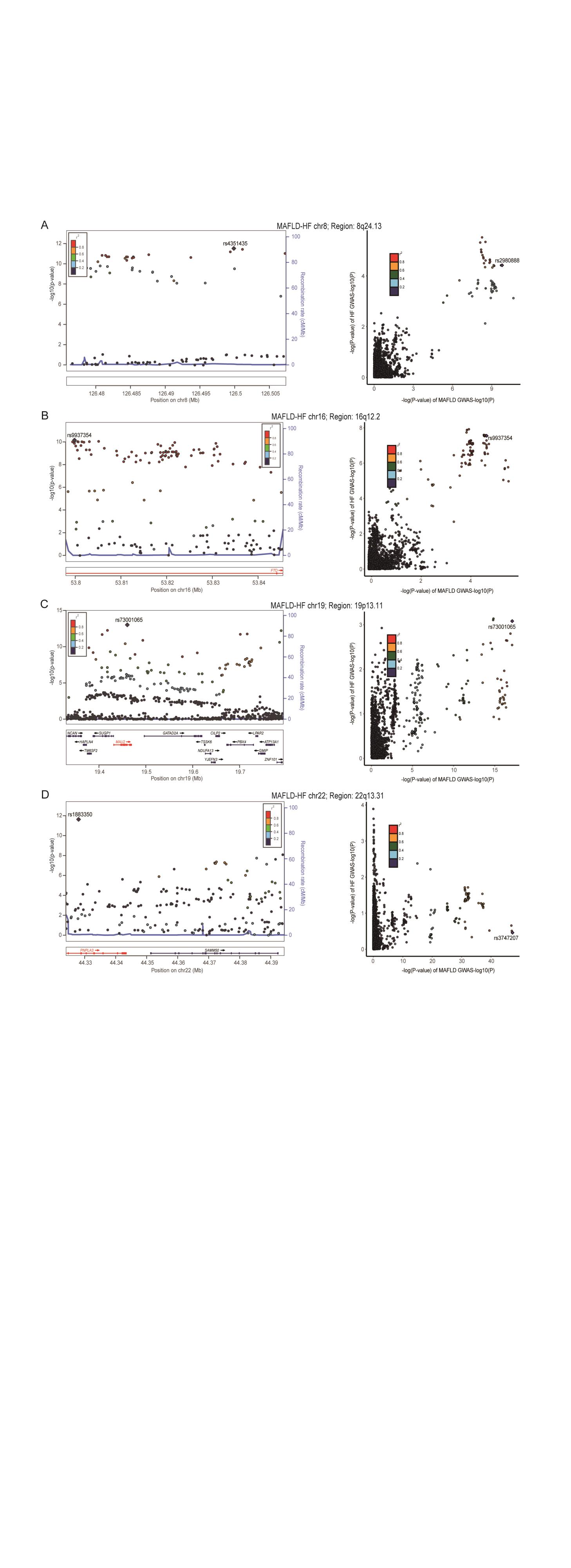

**Supplementary Fig. 4. Locus comparing plots for the shared causal variant for the associations of metabolic dysfunction-associated fatty liver disease and heart failure.**

A total of 49 genomic loci were identified to have strong evidence between MAFLD and CVDs (PP.H4 > 0.7). The left panel depicts the PLACO results using LocusZoom plots, and the right panel compares the two single-trait GWAS statistics for each variant-trait pair using LocusCompare plots. For the LocusZoom plots, the x-axis shows the genomic position of each variant, and the y-axis shows the -log10 P value from the PLCAO results. A purple diamond represents each locus's top variant with the smallest Ppraco. The color of each variant represents its LD relationship with the top variant. For the LocusCompare plots, each dot represents a variant, and the x-axis shows the -log10 PGWAS from the corresponding GWAS for MAFLD, and the y-axis shows the -log10 PGWAS from the corresponding GWAS for CVDs. Purple diamonds also represent candidate-shared causal variants identified by pairwise colocalization analysis. The color of each variant represents its LD relationship with the candidate-shared causal variant. All genomic locations are based on reference genome hg19, and the LD calculation is based on the 1000 Genomes Project of the European population. (A) 8q24.13（rs4351435）in MAFLD-HF, (B) 16q12.2（rs9937354）in MAFLD-HF, (C) 19p13.11（rs73001065）in MAFLD-HF, (D) 22q13.31（rs1883350）in MAFLD-HF. Detailed descriptions were provided in Supplementary Table 7.

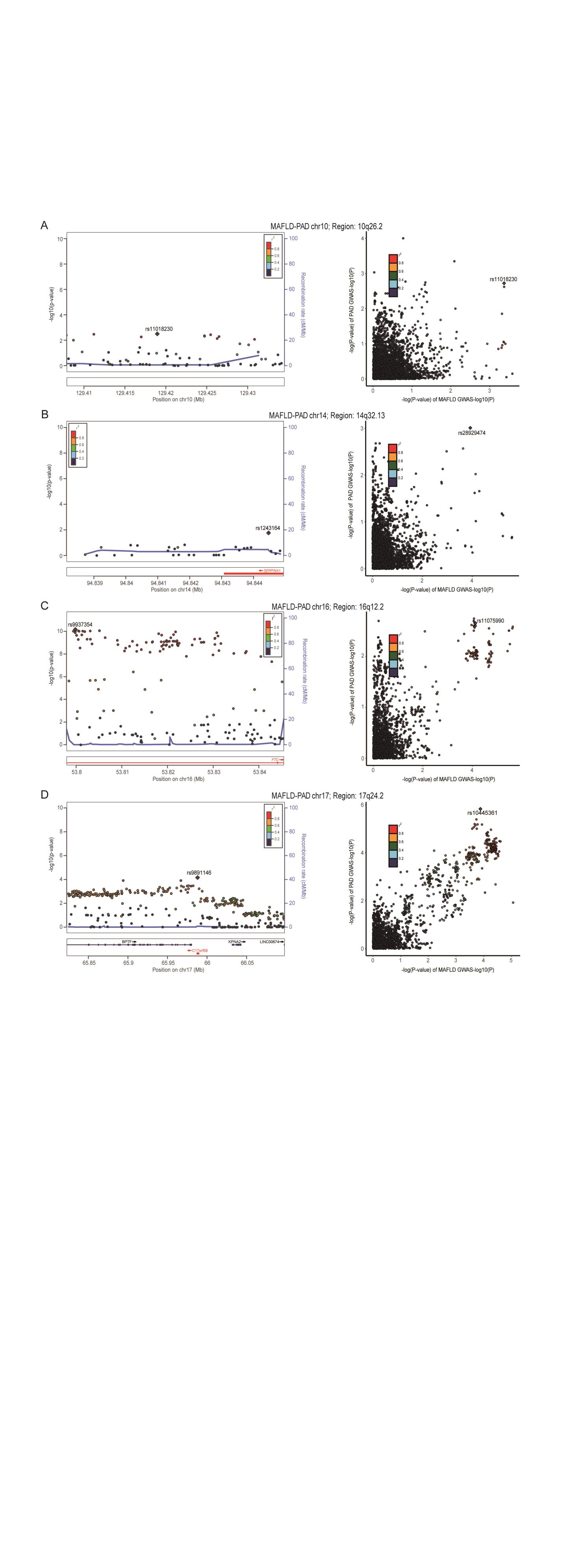

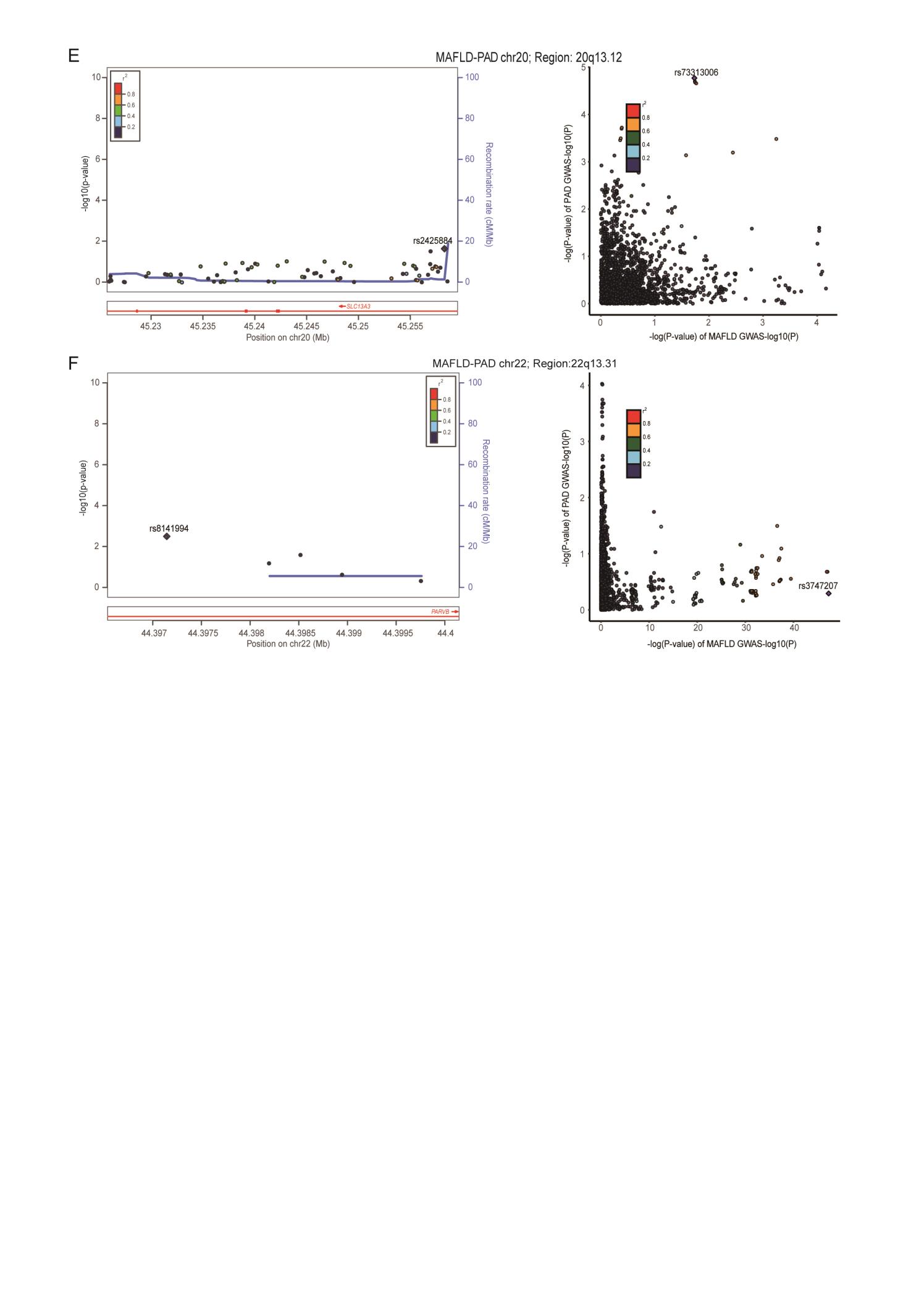

**Supplementary Fig. 5. Locus comparing plots for the shared causal variant for the associations of metabolic dysfunction-associated fatty liver disease and peripheral artery disease.**

A total of 49 genomic loci were identified to have strong evidence between MAFLD and CVDs (PP.H4 > 0.7). The left panel depicts the PLACO results using LocusZoom plots, and the right panel compares the two single-trait GWAS statistics for each variant-trait pair using LocusCompare plots. For the LocusZoom plots, the x-axis shows the genomic position of each variant, and the y-axis shows the -log10 P value from the PLCAO results. A purple diamond represents each locus's top variant with the smallest Ppraco. The color of each variant represents its LD relationship with the top variant. For the LocusCompare plots, each dot represents a variant, and the x-axis shows the -log10 PGWAS from the corresponding GWAS for MAFLD, and the y-axis shows the -log10 PGWAS from the corresponding GWAS for CVDs. Purple diamonds also represent candidate-shared causal variants identified by pairwise colocalization analysis. The color of each variant represents its LD relationship with the candidate-shared causal variant. All genomic locations are based on reference genome hg19, and the LD calculation is based on the 1000 Genomes Project of the European population. (A) 10q26.2（rs11018230）in MAFLD-PAD, (B) 14q32.13（rs28929474）in MAFLD-PAD, (C) 16q12.2（rs11075990）in MAFLD-PAD, (D) 17q24.2（rs10445361）in MAFLD-PAD, (E) 20q13.12（rs117068934）in MAFLD-PAD, (F) 22q13.31（rs13055235）in MAFLD-PAD. Detailed descriptions were provided in Supplementary Table 7.

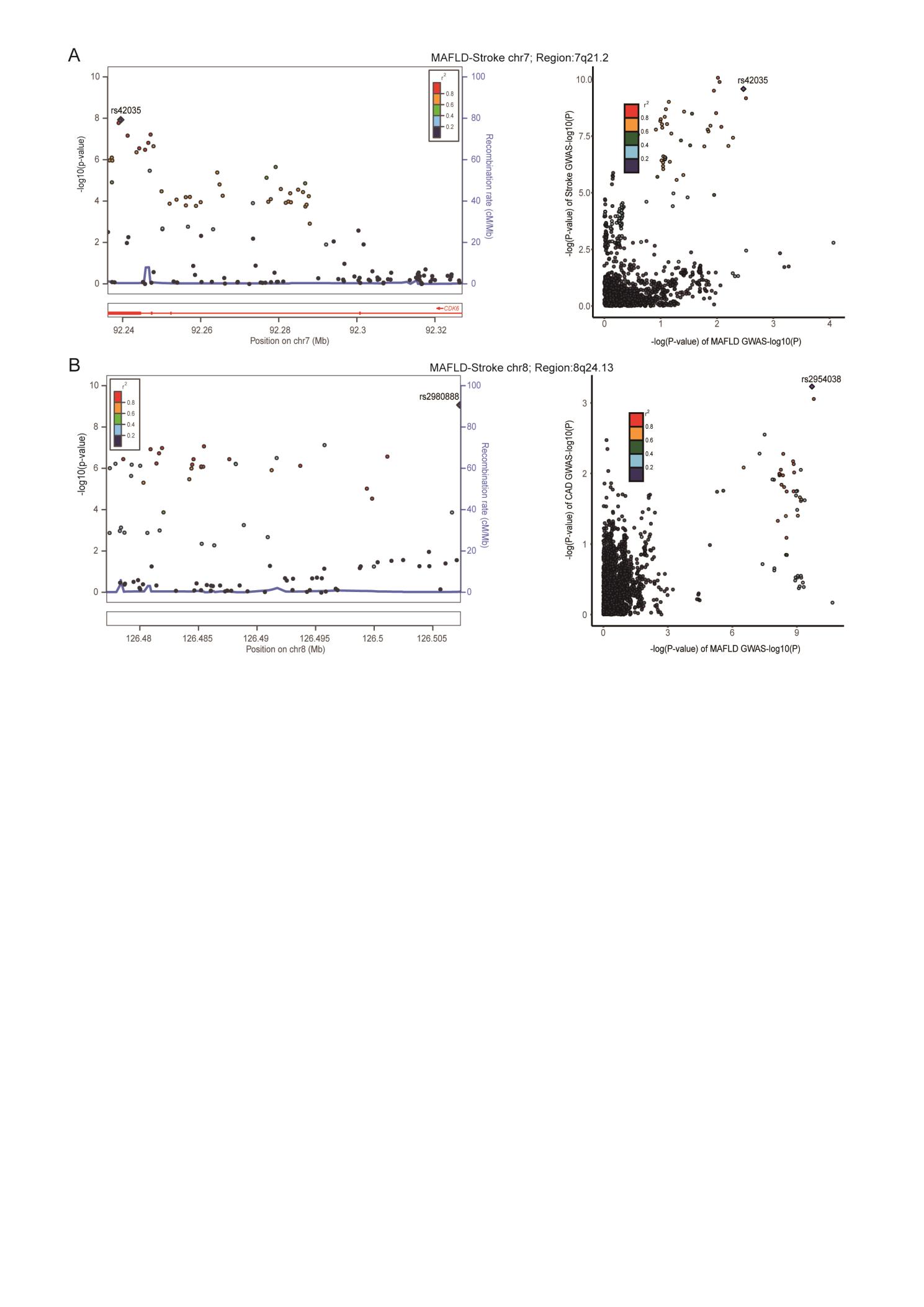

**Supplementary Fig. 6. Locus comparing plots for the shared causal variant for the associations of metabolic dysfunction-associated fatty liver disease and stroke.**

A total of 49 genomic loci were identified to have strong evidence between MAFLD and CVDs (PP.H4 > 0.7). The left panel depicts the PLACO results using LocusZoom plots, and the right panel compares the two single-trait GWAS statistics for each variant-trait pair using LocusCompare plots. For the LocusZoom plots, the x-axis shows the genomic position of each variant, and the y-axis shows the -log10 P value from the PLCAO results. A purple diamond represents each locus's top variant with the smallest Ppraco. The color of each variant represents its LD relationship with the top variant. For the LocusCompare plots, each dot represents a variant, and the x-axis shows the -log10 PGWAS from the corresponding GWAS for MAFLD, and the y-axis shows the -log10 PGWAS from the corresponding GWAS for CVDs. Purple diamonds also represent candidate-shared causal variants identified by pairwise colocalization analysis. The color of each variant represents its LD relationship with the candidate-shared causal variant. All genomic locations are based on reference genome hg19, and the LD calculation is based on the 1000 Genomes Project of the European population. (A) 7q21.2（rs42035）in MAFLD-Stroke, (B) 8q24.13（rs2954038）in MAFLD-Stroke. Detailed descriptions were provided in Supplementary Table 7.
